# Views on Race, Ethnicity, and Algorithmic Reform in Medicine: A Multinational Opt-In Survey

**DOI:** 10.64898/2026.05.15.26352687

**Authors:** Amin Adibi, Xingyu Le, Emma Pierson, James Allen Diao, Nazafarin Esfandiari, Chris Carlsten, Mohsen Sadatsafavi

**Affiliations:** University of British Columbia; UC Berkeley; Harvard Medical School, Brigham and Women’s Hospital

**Keywords:** race and ethnicity, clinical algorithms, algorithmic reform, algorithmic fairness, public opinion, survey

## Abstract

Several professional medical societies have removed race and ethnicity from widely used clinical algorithms affecting millions of patients, yet patient and public opinion on these changes has not been systematically explored. Using four guideline-referenced algorithms representing different approaches to reform, we conducted a cross-sectional, multinational, opt-in survey with quota sampling for equal participation across ten categories of self-identified race and ethnicity. Of 1,050 responses, 994 from 43 countries were eligible (median age 32). Across spirometry, kidney, and cardiovascular calculators, 40.0–47.6% of respondents preferred the race-specific versions and only 16.7–28.2% the race-free alternatives, while 22.0–43.0% felt unrepresented by the categories offered. Respondents expressed greater comfort with race and ethnicity compared to area-based proxies such as postal or ZIP code (75.3% vs. 47.1%), and only 12.8% agreed race and ethnicity should never be used clinically. Findings were largely consistent across countries, self-identified race and ethnicity, and among participants reporting prior experiences of racism in healthcare.

**Data and Code Availability:** A deidentified dataset is available on Zenodo (Adibi et al., 2026). The fully reproducible Quarto source for this paper is available at https://github.com/aminadibi/algorithmic-reform-survey.

**Institutional Review Board (IRB):** The study was approved by the Behavioural Research Ethics Board at the University of British Columbia (H25-02789). All participants gave informed consent at the beginning and at the end of the survey.

## 1. Introduction

Since 2019, a movement has emerged to reconsider the use of race and ethnicity in clinical equations and algorithms (Vyas et al., 2020; Nkinsi and Young, 2022). As of April 2026, several professional societies have removed race as an explicit input from algorithms used to estimate spirometry reference standards^1^ (Bhakta et al., 2023; Bowerman et al., 2023), kidney function^2^ (Delgado et al., 2022), cardiovascular risk for treating dyslipidemia and hypertension^3^ (Khan et al., 2024; Jones et al., 2025; Blumenthal et al., 2026), and risk of vaginal birth after caesarean (American College of Obstetricians and Gynecologists, 2021). Others, including the International Osteoporosis Foundation^4^ and the Society of Thoracic Surgeons, have resisted such calls, citing unacceptable accuracy loss (MacGillivray, 2023; Shahian et al., 2022), particularly for minority groups in the US (Kanis et al., 2024). These revisions affect tens of millions of patients in the US and potentially many more globally (Diao et al., 2024b,a), yet only the kidney function task force reported systematically seeking input from patients and community members (Delgado et al., 2022).

A critical gap in this debate is the relative absence of global patient and public perspectives (Bhakta et al., 2023). Existing opinion studies focus on the US (Schmidt et al., 2023; Jain et al., 2023; Diao et al., 2026), even though these algorithms are deployed globally (Braun, 2014), and the broader debate has likewise been US-centric (Chellappoo, 2024; Doh et al., 2025; Jaime and Kern, 2024). We report a multinational survey built around four guideline-referenced algorithms spanning four distinct reform strategies (Table 1), with opt-in participants in 43 countries across six continents, quota-sampled for equal participation by sex and by ten self-identified categories of race and ethnicity. Beyond measuring preferences, we test whether the race and ethnicity variable itself is stable: we asked each participant to self-classify under several algorithms’ category schemes and measured how often those classifications agreed.

**Table 1:** The four guideline-referenced algorithms presented to participants, their race and ethnicity categories, and their approach to reform. Full details are in Table 2.

| **Original** | **Categories** | **Revised** | **Reform** |
|---|---|---|---|
| 2009 CKD-EPI | Black, non-Black | 2021 CKD-EPI | Refit without race |
| GLI-2012 | White, African American, Northeast Asian, Southeast Asian, other<sup>1</sup> | GLI-Global | Race-averaged |
| Pooled Cohort | White, African American, other<sup>2</sup> | PREVENT | Race replaced by optional ZIP code |
| FRAX | Country-specific<sup>3</sup> | — | Race retained |

<sup>1</sup>The average of the four group-specific equations.
<sup>2</sup>In the web calculator only, where it returns the White value.
<sup>3</sup>Ethnicity-specific in five countries; a single model elsewhere.

**Table 2:**
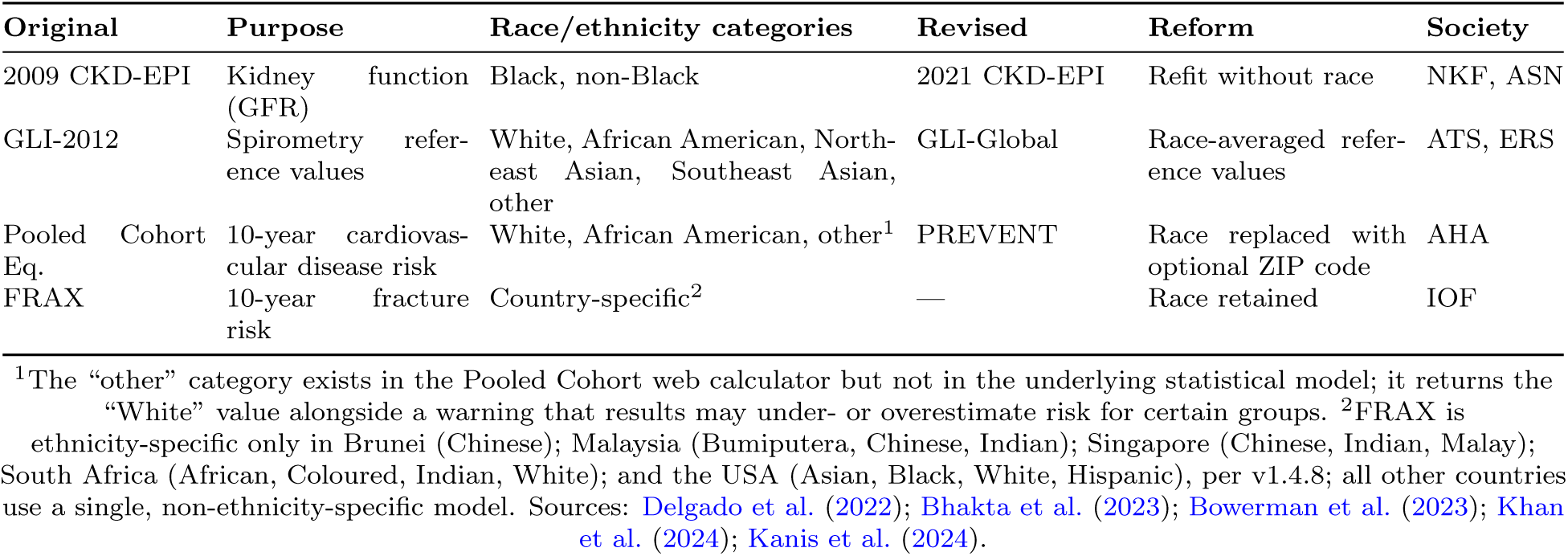
The four guideline-referenced algorithms presented to participants, with the race and ethnicity category scheme each one offers.

| **Original** | **Purpose** | **Race/ethnicity categories** | **Revised** | **Reform** | **Society** |
|---|---|---|---|---|---|
| 2009 CKD-EPI | Kidney function (GFR) | Black, non-Black | 2021 CKD-EPI | Refit without race | NKF, ASN |
| GLI-2012 | Spirometry refer-ence values | White, African American, North-east Asian, Southeast Asian, other | GLI-Global | Race-averaged refer-ence values | ATS, ERS |
| Pooled Cohort Eq. | 10-year cardiovas-cular disease risk | White, African American, other<sup>1</sup> | PREVENT | Race replaced with optional ZIP code | AHA |
| FRAX | 10-year fracture risk | Country-specific<sup>2</sup> | — | Race retained | IOF |

<sup>1</sup>The “other” category exists in the Pooled Cohort web calculator but not in the underlying statistical model; it returns the “White” value alongside a warning that results may under- or overestimate risk for certain groups.
<sup>2</sup>FRAX is ethnicity-specific only in Brunei (Chinese); Malaysia (Bumiputera, Chinese, Indian); Singapore (Chinese, Indian, Malay); South Africa (African, Coloured, Indian, White); and the USA (Asian, Black, White, Hispanic), per v1.4.8; all other countries use a single, non-ethnicity-specific model. Sources: Delgado et al. (2022); Bhakta et al. (2023); Bowerman et al. (2023); Khan et al. (2024); Kanis et al. (2024).

## 2. Methods

The questionnaire was drafted iteratively with feedback from six graduate students and three faculty members spanning epidemiology, clinical prediction modelling, medicine, and algorithmic fairness, then revised after cognitive interviews with two patient partners. Full wording is provided as supplementary material.

After a plain-language explainer, the questionnaire presented the four algorithms of Table 1 in randomized order. For each, participants self-classified under that algorithm’s category scheme and rated, on 5-point Likert items with randomized response direction, whether they felt represented by its categories, whether using race or ethnicity could improve their care, and whether they worried it might cause harm. After each section, participants saw the race-free version, where one exists, and chose which they preferred. Comfort with individual predictors was assessed via a hypothetical cancer risk calculator covering postal or ZIP code, race or ethnicity, genetic ancestry, biological sex, country of origin, age, and environmental exposures. Further items covered experiences of unfair treatment in healthcare, whether staff had recorded race or ethnicity without asking, and four items replicating a prior US survey (Diao et al., 2026).

Participants were recruited through Prolific with quota sampling for equal participation by sex as recorded on legal/official documents and across ten self-identified ethnicity groups previously collected by the platform. The survey was distributed over four waves at midnight and noon Pacific Time in January 2026. To expand geographical diversity, residents of countries exceeding 100 responses were excluded from subsequent waves. Participants consented at the beginning and at the end of the survey; those who completed the study and passed authenticity checks were reimbursed at an average rate of £9.72/hour. Self-identified data on age, sex, gender, ethnicity, countries of residence and birth, employment, and socioeconomic status were collected by the platform prior to the survey. Because the platform’s ethnicity field required a single category, an additional survey item asked participants to select all population groups that applied, or to self-describe, with options including African, African American, Caribbean, East Asian, Hispanic/Latino, Indigenous, Middle Eastern and North African, South Asian, Southeast Asian, and White.

Quantitative analyses were descriptive and unweighted (Quarto v1.9.30, R v4.5.2), with 95% confidence intervals from 2,000 nonparametric bootstrap resamples; open-ended responses were analyzed by conventional content analysis (Hsieh and Shannon, 2005) using a double-coded shared codebook (pooled inter-rater agreement 94.9%, Cohen’s *κ* 0.79, Gwet’s AC1 0.93; Appendix H; example quotes in Appendix I). We define *net comfort* as the percentage extremely or somewhat comfortable minus the percentage extremely or somewhat uncomfortable, and *net agreement* analogously. The study followed AAPOR best practices; further detail is in Appendix A.

## 3. Results

Of 1,050 responses, 48 participants revoked consent, 6 timed out, and 2 failed authenticity checks. The remaining 994 lived in 43 countries, were born in 87, and had a median age of 32.0 years (IQR 26–41); 500 (50.3%) were female and 494 (49.7%) male by sex as recorded on legal documents, and 47.7% worked full-time (Table 4). No survey-collected variable was missing for more than 0.7% of participants. Pooled percentages below describe this recruited sample and are not population-weighted estimates.

### Comfort with predictors

Respondents were most comfortable with age (net comfort +92.3%, 95% CI +90.1 to +94.4), biological sex (+89.4%, +86.9 to +91.7), environmental exposures (+88.7%, +85.9 to +91.2), and genetic ancestry (+82.2%, +78.9 to +85.2). Comfort declined for country of origin (+68.8%, +64.8 to +72.7) and race and ethnicity (+62.4%, +57.8 to +66.9), and fell sharply for ZIP or postal code (+14.5%, +9.1 to +20.0; Figure 1). Net comfort with race and ethnicity stayed positive in every self-identified group and in every country with *≥*20 respondents (+44.6% among Africans to +77.7% among East Asians), whereas comfort with ZIP or postal code was negative in several (Appendix E). Only 12.8% (10.7–14.9) agreed doctors should never use race or ethnicity, from 7.6% (4.4–11.1) among White to 22.2% (11.1–33.3) among African American respondents, whereas 87.9% (85.8–89.9) were comfortable with its use if the doctor explained why and how.

### Reactions to reform

Across all four race-specific tools (Figure 2), 44.8–64.8% of participants felt represented by the categories offered (while 30.9–42.9% felt unrepresented across the three reformed calculators and 22.1% for FRAX), with net agreement ranging from +1.8% (*−*4.2 to +7.7) for the binary Pooled Cohort Equations to +42.8% (+37.6 to +47.8) for the country-specific FRAX. A majority also believed incorporating race helped improve their care (55.5– 58.0%; net +33.9% to +40.1%), and roughly half disagreed that it could cause harm (net *−*21.2% to *−*28.5%).

Respondents did not uniformly embrace the revised algorithms. For spirometry and eGFR, 40.0% (36.9–43.2) and 40.0% (36.9–43.1) preferred the race-based versions, versus 28.2% (25.3–31.1) and 26.9% (24.1–29.6) for the revised replacements. For cardio-vascular risk, 47.6% (44.4–50.6) preferred the race-based Pooled Cohort Equations over 16.7% (14.5– 19.0) preferring PREVENT, which substitutes optional ZIP code for race. The remaining 31.8% to 35.7% were neutral, unsure, or self-described their preference across the three pairs. Asked whether concerns differed between AI systems and simpler formula-based tools, nearly three times as many respondents were more concerned about AI (24.1%) than less (8.7%), though most drew no distinction (Appendix B).

### Categories are not stable

Self-identification shifted with the categories each algorithm offered (Figure 3). Multiethnic participants and populations excluded from algorithm categories were most likely to classify themselves inconsistently with developers’ intent. Only 51.7% (48.8–54.7) chose the developer-intended category in all three algorithms, ranging from 57.7% (54.6–60.9) for GLI-2012 and 69.2% (66.4–72.1) for the Pooled Cohort Equations to 90.1% (88.2–92.0) for eGFR. Agreement in all three ranged from 92.7% among White to 22.1% among East Asian participants, and was lowest for groups lacking a dedicated category in one or more algorithms (Table 3). Although the GLI-2012 developers treated Middle Eastern and Hispanic individuals as part of the White category (Quanjer et al., 2012), only 40.6% (31.2–51.0) of Middle Eastern and 37.5% (28.8–47.1) of Hispanic/Latino respondents selected White when offered the GLI-2012 scheme. Separately, 21.4% (18.8–24.0) reported staff recording their race or ethnicity without asking, and 5.4% of all participants were misidentified as a result.

**Figure 1:**
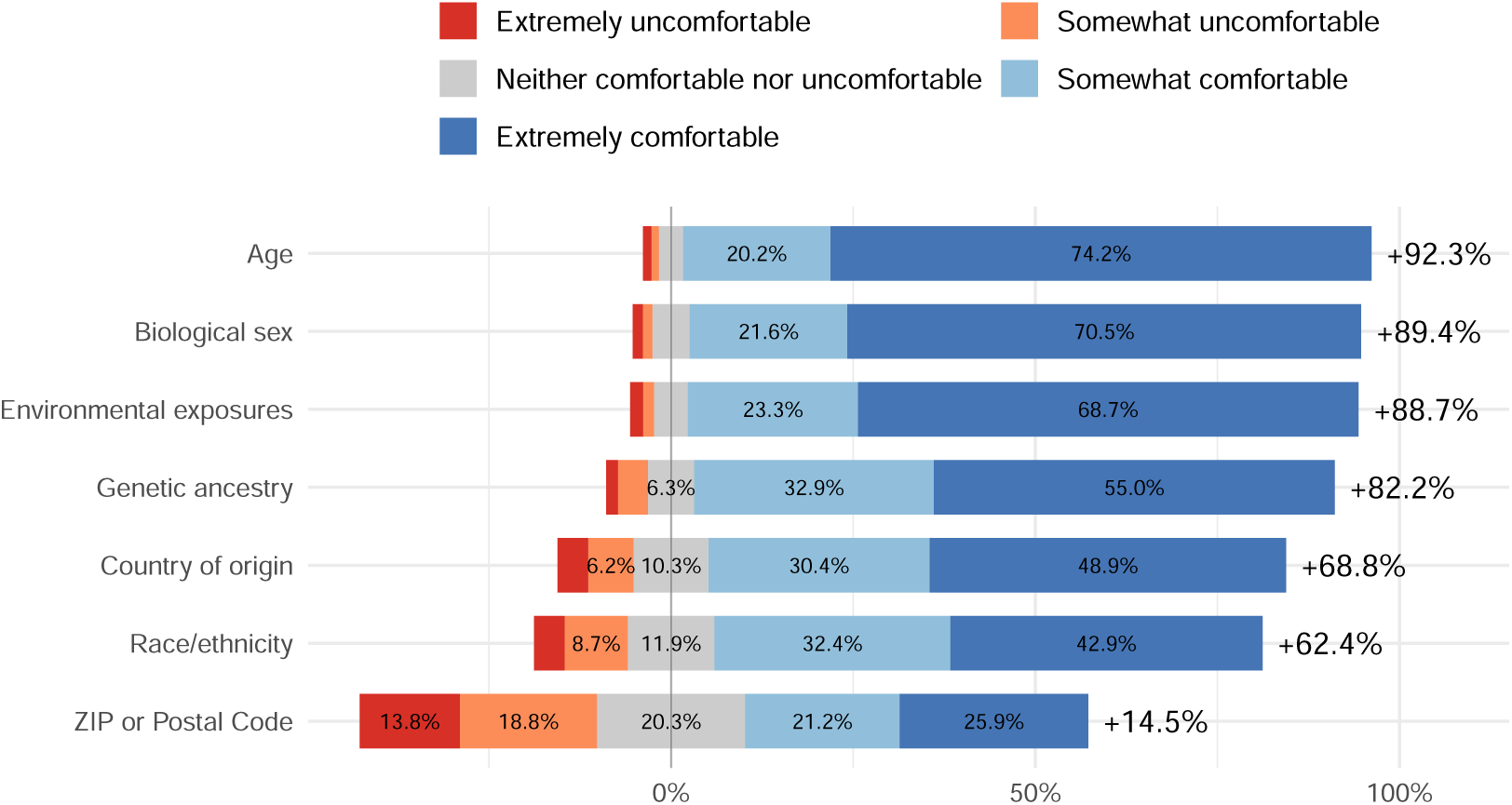
Comfort with each factor in a hypothetical cancer risk calculator. Bars extend rightward for comfortable and leftward for uncomfortable responses, with neutral responses (grey) centred at zero. Ordered by net comfort.

### Participants who had experienced racism

Overall, 19.8% (17.4–22.1) reported unfair healthcare treatment because of their race or ethnicity, from 4.9% among White to 41.8% (29.1–54.5) among African Americans, and 54.1% viewed racial bias as a major problem in healthcare. This subgroup reported lower net comfort with every predictor—most for race and ethnicity (+41.6% vs. +62.4%)—and felt less represented by the GLI-2012 and FRAX categories (Appendix D). Most notably, while the full sample disagreed that race-specific algorithms could cause harm (net *−*21.2% to *−*28.5%), this subgroup shifted toward agreement (net *−*3.0% to +6.6%). Even so, a majority still agreed that using race helps improve care, and a plurality still preferred the race-specific version—except for spirometry, where 40.1% (33.5–46.7) preferred race-averaged GLI-Global over 35.5% (28.9–42.6) for GLI-2012.

## 4. Discussion

Our sample is not representative, but it spans a wider range of cultures and geographies than prior work on this question, and it yields four findings. First, most respondents were comfortable with race or ethnicity as a predictor in clinical calculators (75.3%), consistently across countries, self-identified groups, and among those who had experienced racism in healthcare. Second, across spirometry, eGFR, and cardio-vascular risk, 40.0–47.6% preferred the race-specific versions and only 16.7–28.2% the race-free alternatives. Third, a substantial minority did not see themselves in the categories offered, from 22.1% for FRAX to 42.9% for the Pooled Cohort Equations. Fourth, 2.5 times as many respondents were uncomfortable with postal or ZIP code as with race or ethnicity, even though ZIP code has been adopted as a replacement in some revised algorithms.

The instability we observed has direct consequences for machine learning in health. A covariate whose value depends on which category set the interface happens to offer is a noisy and non-transportable input, and any fairness audit stratified on it inherits that noise: the same person may enter one subgroup under GLI-2012 and another under the Pooled Cohort Equations. Although many respondents described race and ethnicity in biological terms, their own self-classification was fluid. These are socially determined constructs (Bonilla-Silva, 1999) with complex biological associations (Rose, 2023; Holland, 2008), and our results are consistent with evidence that recorded race varies with who observes it (Roth, 2016; Herman, 2010), when (Liebler et al., 2017), social status cues (Freeman et al., 2011; Saperstein et al., 2014), and the classification scheme (Nicolas et al., 2019; Harris and Sim, 2002), sometimes with measurable consequences for model performance (Movva et al., 2023; Adibi et al., 2025a).

**Figure 2:**
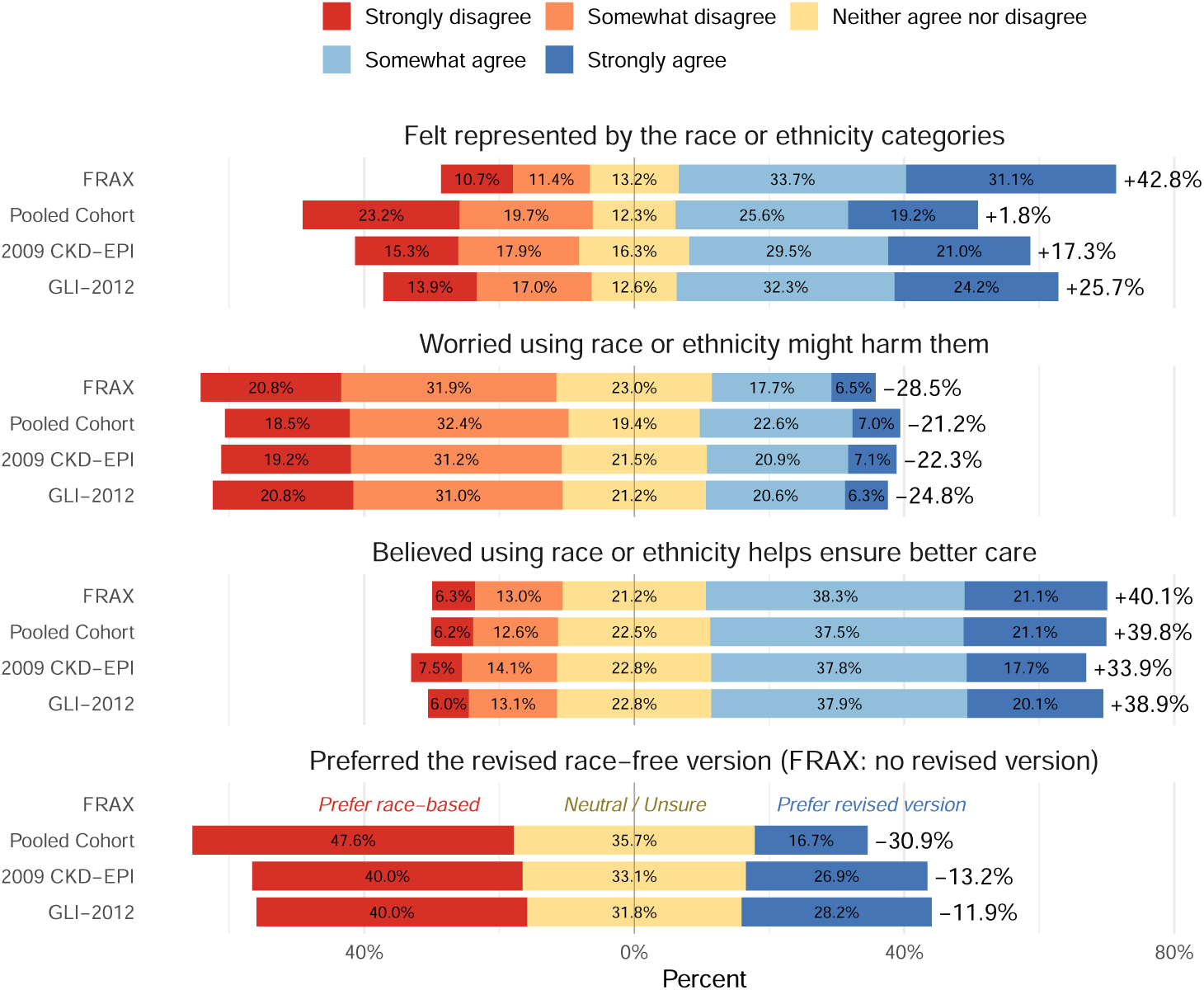
Reactions to race-specific algorithms and preference for their revised counterparts. Upper panels: agreement with three statements per algorithm. Lower panel: preference for the revised race-free version. Bars extend rightward for agreement or for the revised version and leftward for the race-based version; neutral (yellow) centred at zero.

The discomfort with ZIP or postal code suggests that swapping race for an area-based deprivation score may not read as more trustworthy to patients (Adibi et al., 2025b), even though at least three widely used cardiovascular calculators— QRISK3, PREVENT, and ASSIGN (Hippisley-Cox et al., 2017; Khan et al., 2024; Woodward et al., 2007)—take these codes as an optional input to look up deprivation indices; how often patients are told remains unstudied. Our findings also suggest a gap between public and professional views: in a US survey of 66 pulmonologists, 64% recommended race-averaged spirometry references versus 9% recommending the race-specific version (Brems et al., 2026). Our results align with the one prior representative US survey, in which 79.8% of 1,750 adults preferred their physician use all available information, including race (Diao et al., 2026); our sample was younger, more diverse, and more likely to view racism as a major problem in healthcare, yet was still less likely to say doctors should never use race (12.8% vs. 22.0%) and more uncomfortable with postal or ZIP code (32.6% vs. 22.3%). None of this implies race-free algorithms are fairer or less fair: algorithms can be biased regardless of whether they include race (Obermeyer et al., 2019; Pierson et al., 2021; Zink et al., 2024; Siddique et al., 2024).

**Figure 3:**
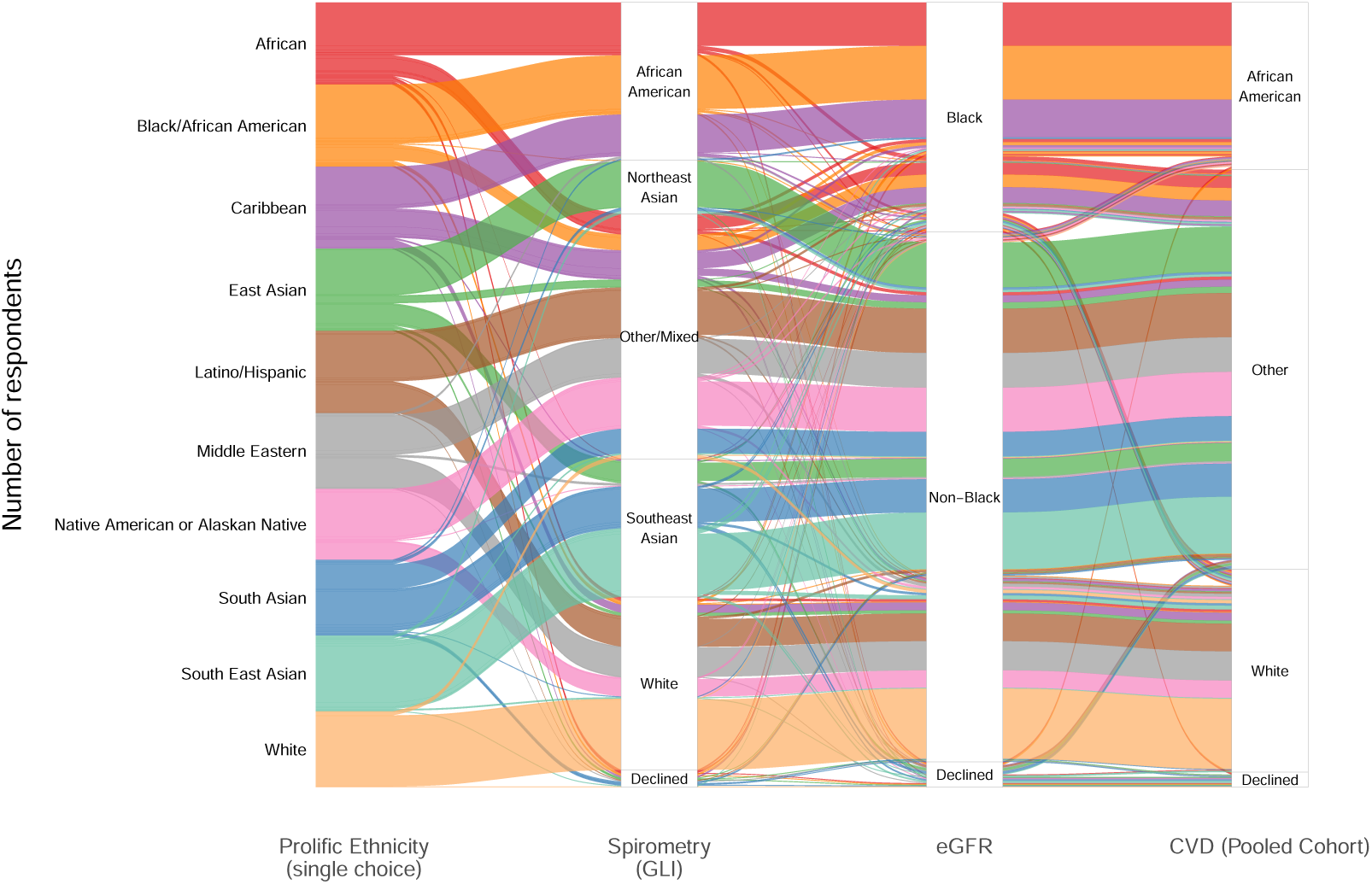
Racial and ethnic self-identification shifts as the available categories change across classification systems used in clinical calculators. Each vertical bar is one system: survey ethnicity (the single-choice category selected at platform sign-up), then the categories offered by spirometry references (GLI), eGFR, and cardiovascular risk (Pooled Cohort). Each ribbon represents respondents sharing the same combination of choices across all four systems, with width proportional to the number of respondents on that path. Ribbons that fan out or merge across columns mark respondents who did not carry the same label from one system to the next. For example, GLI-2012 offers no dedicated Middle Eastern or Hispanic/Latino option, yet only 40.6% of Middle Eastern and 37.5% of Hispanic/Latino respondents chose ‘White’, the category the developers had intended for them.

### Limitations

These are initial reactions: respondents were not primed with arguments or evidence and might shift given more information. The design was not intended to yield representative samples of individual countries. Country-level sample sizes varied substantially. Country-specific observations should therefore be interpreted cautiously. The Prolific pool skews younger and more educated and excludes populous nations such as China and Pakistan. Self-selection by topic, reward rate, and survey length may introduce hard-to-quantify response biases (Aksoy and Nevo, 2025). While the questionnaire randomized the order of algorithms and the direction of Likert items, race-specific algorithms always appeared before their revised versions to mirror the clinical implementation. We cannot rule out the possibility that this ordering might have influenced responses.

## Conclusion

Choosing which factors enter a clinical algorithm is not a purely technical decision. Philosophers of science have long argued that researchers cannot avoid value judgments under uncertainty (Rudner, 1953; Douglas, 2000), and that the values informing them should reflect public rather than expert values alone (Douglas, 2005; Kitcher, 2011)—an argument sharpened by the history of racism in healthcare (National Academies of Sciences, Engineering, and Medicine, 2025) and by evidence that scientists are better served by genuine dialogue than by top-down communication (Cologna et al., 2025). The lack of widespread community consultation in algorithmic reform is a missed opportunity to raise awareness, align values, and preserve public trust. Professional societies and algorithm developers should engage patient and public opinion, whether or not their final recommendation is to remove race and ethnicity as predictors.

## Supporting information

Questionnaire

## Data Availability

Data required for reproducing results is available online at https://zenodo.org/records/20403996

https://github.com/aminadibi/algorithmic-reform-survey

## Acknowledgments

The study was funded by the University of British Columbia Public Scholars Initiative and Legacy for Airway Health. AA is also supported by a University of British Columbia 4-Year Doctoral Fellowship and RESPNet BC. We thank our patient partners for sharing their feedback and insights throughout this study, and are grateful to Legacy for Airway Health, Dr. Phalgun Joshi, and Beth White for supporting patient partner engagement. We are also grateful to Dr. Kennedy Borle and Dr. Alexa Norton for their advice on ethics application and study design. The authors acknowledge that most of the activities of this research project were conducted on the traditional, ancestral, and unceded territories of the Musqueam and Kwikwetlem Nations.

## Appendix A. Extended Methods

### Questionnaire design

The questionnaire was iteratively drafted with feedback from six graduate students (epidemiology, health outcomes, clinical prediction modelling) and three faculty members (medicine, clinical prediction, algorithmic fairness), then revised after pretesting via cognitive interviews with two patient partners. The exact wording of every item is provided in the supplementary material.

The four reform strategies presented were: refitting without race (eGFR), race-averaged reference values (spirometry), replacing race with an area-based deprivation score derived from postal codes (PREVENT), and retaining race or ethnicity (FRAX). The three Likert items per algorithm were phrased as *Represented* (“I feel represented by the race or ethnicity categories used to [interpret lung function test results / estimate kidney function / estimate heart disease risk / estimate bone fracture risk]”), *Worry harm* (“I worry that using my race or ethnicity in [algorithm] to support clinical decisions might harm me”), and *Helps care* (“I believe that using race or ethnicity in [algorithm] could help ensure better care for me”).

Participants were asked whether they had ever felt treated unfairly in the healthcare system because of their race or ethnicity, with an optional open-ended follow-up, and whether clinic staff had ever recorded their race or ethnicity without asking. Four replication items enabled comparison with prior representative US surveys (Diao et al., 2026), covering whether doctors should use race or ethnicity in clinical care, perceptions of bias in health and medicine, and expectations of care from racially or ethnically concordant providers.

### Analysis

Open-ended responses were analyzed by conventional content analysis (Hsieh and Shannon, 2005) in NVivo v1.7.2; non-English responses were translated with Google Translate and cross-checked with DeepL. Two reviewers developed a shared codebook through open coding of a subset of responses, then independently applied it to 10% of responses per question to assess consistency. We report percent agreement, Cohen’s *κ*, and Gwet’s AC1 over all response-by-code decisions, with 95% percentile bootstrap confidence intervals from 2,000 resamples of responses; AC1 is reported alongside *κ* because *κ* is unstable for rarely applied codes. After discrepancies were resolved by consensus, the remaining responses were coded by one reviewer. Codes were grouped into broader categories by conceptual similarity, frequencies tallied, and representative quotes selected. Quantitative analyses were descriptive and unweighted, performed in Quarto v1.9.30 and R v4.5.2, with 95% confidence intervals from 2,000 nonparametric bootstrap resamples. Agreement with developer-intended categories was defined, for each algorithm, as selecting the category its developers intended for the participant’s survey ethnicity (e.g., White for Middle Eastern and Hispanic/Latino participants in GLI-2012 (Quanjer et al., 2012)); “Prefer not to answer” was counted as disagreement.

## Appendix B. Extended Results

### What race and ethnicity mean to participants

Of 993 substantive responses (three translated from Portuguese, Polish, and Spanish before non-mutually-exclusive coding), many defined race and ethnicity through physical traits and skin colour (37.3%), cultural or historical background (29.5%), place of origin (26.8%), ancestry (22.8%), and genetics (17.0%), while few described them as social constructs (2.9%). About 12.4% explicitly distinguished the two concepts, race relating to visible physical characteristics and ethnicity to cultural background or upbringing. Some (7.0%) described race and ethnicity as central to their sense of community, providing togetherness, belonging, and pride, and 4.5% noted perceived differences in disease risk across racial or ethnic groups (Appendix I).

### Comfort by subgroup

Net comfort with race and ethnicity ranged from +44.6% (95% CI +31.5 to +57.7) among Africans to +77.7% (+66.0 to +88.3) among East Asians, and for ZIP or postal code from *−*13.4% (*−*30.4 to +3.6) among Hispanic/Latino respondents to +30.5% (+13.3 to +47.6) among South Asians. Among countries with *≥*20 respondents, net comfort remained positive for race and ethnicity but was sometimes negative for ZIP or postal code. Overall, 87.9% (85.8–89.9) were comfortable with race or ethnicity in clinical algorithms if the doctor explained why and how it was used, 80.4% (77.9–82.8) if recommended by guidelines, and 79.0% (76.3–81.6) if the doctor believed it best for care.

### Being asked, and being misidentified

About 29.5% (26.8–32.2) recalled a clinician telling them their race or ethnicity played a role in a medical decision. Asked how often they are asked for race or ethnicity in healthcare settings, 8.8% said always, 15.2% most of the time, 8.2% about half the time, 34.1% sometimes, and 33.7% never. In this sample, respondents from the UK, US, and South Africa were least likely to report never being asked. Staff recording race or ethnicity without asking was most often reported in Kenya and South Africa, and misidentification in the US, Kenya, and France.

### AI versus formula-based tools

Asked how concerns about race or ethnicity differed between AI systems and simpler formula-based tools, opinions were divided: the largest group, 29.7% (26.8–32.3), expressed no concern about any tool, and a similar proportion, 29.0% (26.2–31.9), were equally concerned regardless of type. Among those who did differentiate, nearly three times as many were more concerned about AI (24.1%, 21.5–26.8) than less concerned (8.7%, 6.9–10.6).

### Open-ended concerns about reform

Among 215 substantive responses, recurring themes were oversimplification of broad categories such as “Black”, “White”, or “Hispanic” (7.0%) and risk of bias or discrimination grounded in historical misuse (11.2%). Many participants would accept race-specific tools conditionally if shown to improve accuracy or outcomes (8.8%) or on a case-by-case basis by disease indication (7.9%). Notably, 17.2% supported using race clinically, reasoning that it carries biological implications or proxies environmental or lifestyle factors, and 2.8% expressed full trust in providers and researchers (Appendix I).

### Accounts of racism in healthcare

About 9.1% of participants shared experiences of racism in healthcare; of 95 responses, 2 were excluded for non-substantive content and 3 for describing settings outside healthcare. The most common accounts involved differential treatment such as poorer provider attitudes or quality of care (30.0%), dismissive or neglectful behaviour (28.9%), and longer wait times or lower priority (22.2%). Participants also reported stereotypical assumptions about pain tolerance, background, lifestyle, or socioeconomic status, and about individuals with mixed backgrounds, particularly among Indigenous respondents. Overall, 54.1% (50.6–57.6) viewed racial bias as a major problem in healthcare, 34.2% (30.8–37.4) as minor, and 11.8% (9.6–14.0) as not a problem.

### Agreement with developer-intended categories

Table 3 reports, for each algorithm, the proportion of participants who selected the category its developers intended for their survey ethnicity. Agreement was highest for the binary eGFR scheme (90.1%) and lowest for GLI-2012 (57.7%); only 51.7% agreed in all three. Agreement in all three was highest among White participants (92.7%) and lowest among East Asian (22.1%), South Asian (32.3%), Latino/Hispanic (33.7%), and Middle Eastern (38.5%) participants, groups with no dedicated category in one or more algorithms.

**Table 3:**
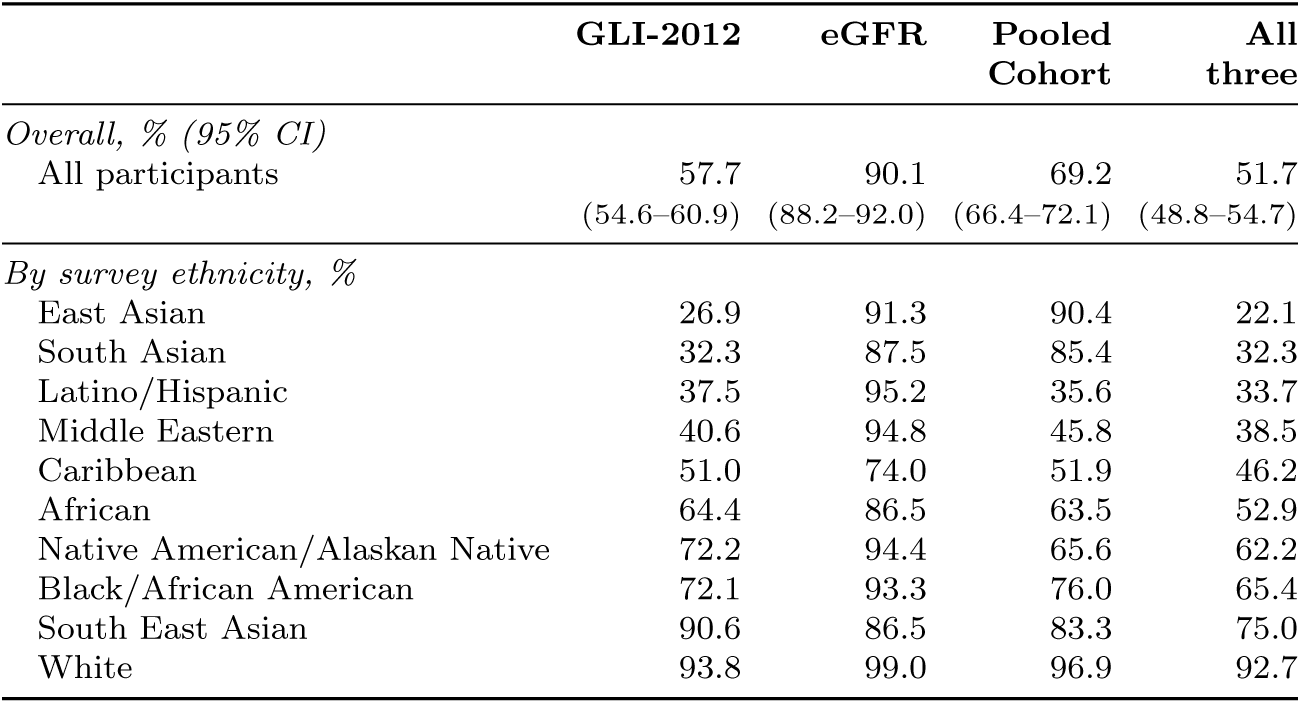
Agreement between participants’ self-classification and the category intended by algorithm developers, overall (% with 95% CI) and by survey ethnicity (%). “Prefer not to answer” was counted as disagreement. Survey ethnicity groups are ordered by agreement in all three algorithms.

|  | **GLI-2012** | **eGFR** | **Pooled Cohort** | **All three** |
|---|---|---|---|---|
| *Overall, % (95% CI)* |  |  |  |  |
| All participants | 57.7<br>(54.6–60.9) | 90.1<br>(88.2–92.0) | 69.2<br>(66.4–72.1) | 51.7<br>(48.8–54.7) |
| *By survey ethnicity, %* |  |  |  |  |
| East Asian | 26.9 | 91.3 | 90.4 | 22.1 |
| South Asian | 32.3 | 87.5 | 85.4 | 32.3 |
| Latino/Hispanic | 37.5 | 95.2 | 35.6 | 33.7 |
| Middle Eastern | 40.6 | 94.8 | 45.8 | 38.5 |
| Caribbean | 51.0 | 74.0 | 51.9 | 46.2 |
| African | 64.4 | 86.5 | 63.5 | 52.9 |
| Native American/Alaskan Native | 72.2 | 94.4 | 65.6 | 62.2 |
| Black/African American | 72.1 | 93.3 | 76.0 | 65.4 |
| South East Asian | 90.6 | 86.5 | 83.3 | 75.0 |
| White | 93.8 | 99.0 | 96.9 | 92.7 |

## Appendix C. Participant Characteristics

Table 4 reports the demographic composition of the analytic sample. Figures 4 and 5 show respondents’ countries of residence and birth. Figure 6 shows how often participants claimed more than one identity, and Figure 7 compares the single category each participant selected at platform sign-up against the multi-select identities they reported in our survey.

**Figure 4:**
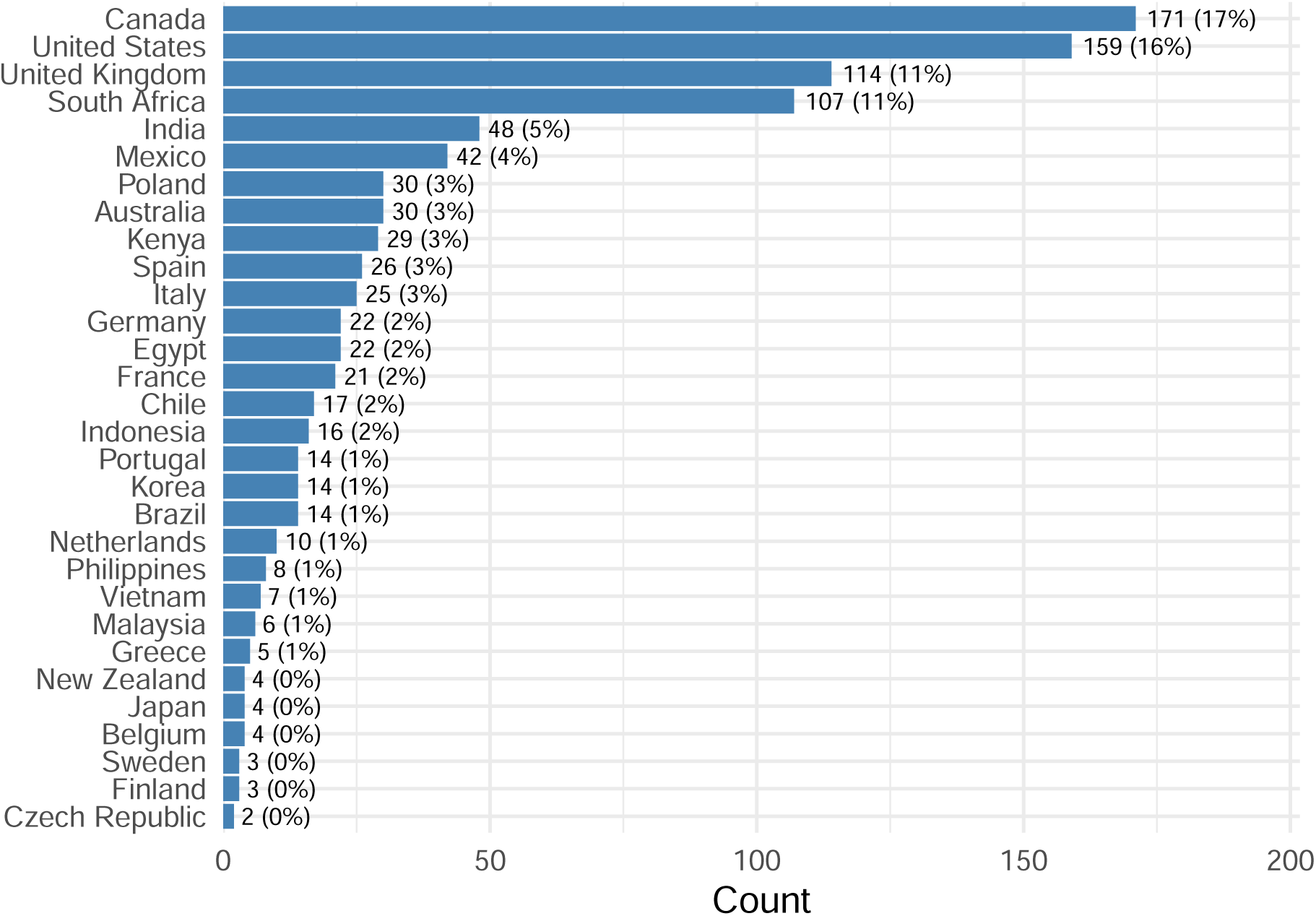
Respondents’ country of residence (top 30).

**Table 4:**
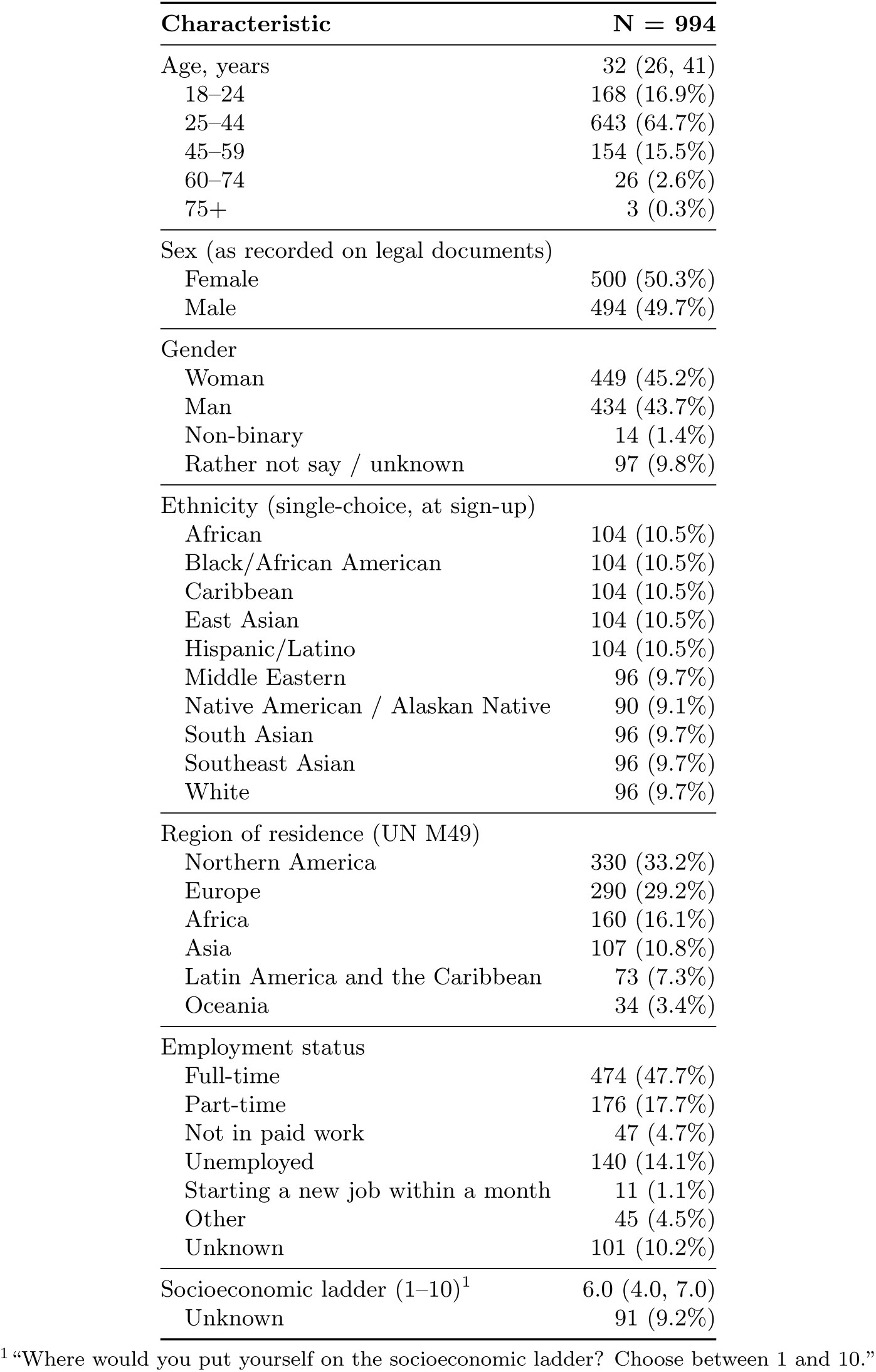
Characteristics of participants. Quota sampling used to obtain approximately equal numbers across sex as recorded on legal documents and ten self-identified groups of race and ethnicity, based on the single-choice category selected at platform sign-up. Values are median (Q1, Q3) or n (%).

| **Characteristic** | **N = 994** |
|---|---|
| Age, years | 32 (26, 41) |
| 18–24 | 168 (16.9%) |
| 25–44 | 643 (64.7%) |
| 45–59 | 154 (15.5%) |
| 60–74 | 26 (2.6%) |
| 75+ | 3 (0.3%) |
| Sex (as recorded on legal documents) |  |
| Female | 500 (50.3%) |
| Male | 494 (49.7%) |
| Gender |  |
| Woman | 449 (45.2%) |
| Man | 434 (43.7%) |
| Non-binary | 14 (1.4%) |
| Rather not say / unknown | 97 (9.8%) |
| Ethnicity (single-choice, at sign-up) |  |
| African | 104 (10.5%) |
| Black/African American | 104 (10.5%) |
| Caribbean | 104 (10.5%) |
| East Asian | 104 (10.5%) |
| Hispanic/Latino | 104 (10.5%) |
| Middle Eastern | 96 (9.7%) |
| Native American / Alaskan Native | 90 (9.1%) |
| South Asian | 96 (9.7%) |
| Southeast Asian | 96 (9.7%) |
| White | 96 (9.7%) |
| Region of residence (UN M49) |  |
| Northern America | 330 (33.2%) |
| Europe | 290 (29.2%) |
| Africa | 160 (16.1%) |
| Asia | 107 (10.8%) |
| Latin America and the Caribbean | 73 (7.3%) |
| Oceania | 34 (3.4%) |
| Employment status |  |
| Full-time | 474 (47.7%) |
| Part-time | 176 (17.7%) |
| Not in paid work | 47 (4.7%) |
| Unemployed | 140 (14.1%) |
| Starting a new job within a month | 11 (1.1%) |
| Other | 45 (4.5%) |
| Unknown | 101 (10.2%) |
| Socioeconomic ladder (1–10)<sup>1</sup> | 6.0 (4.0, 7.0) |
| Unknown | 91 (9.2%) |

<sup>1</sup>“Where would you put yourself on the socioeconomic ladder? Choose between 1 and 10.”

**Figure 5:**
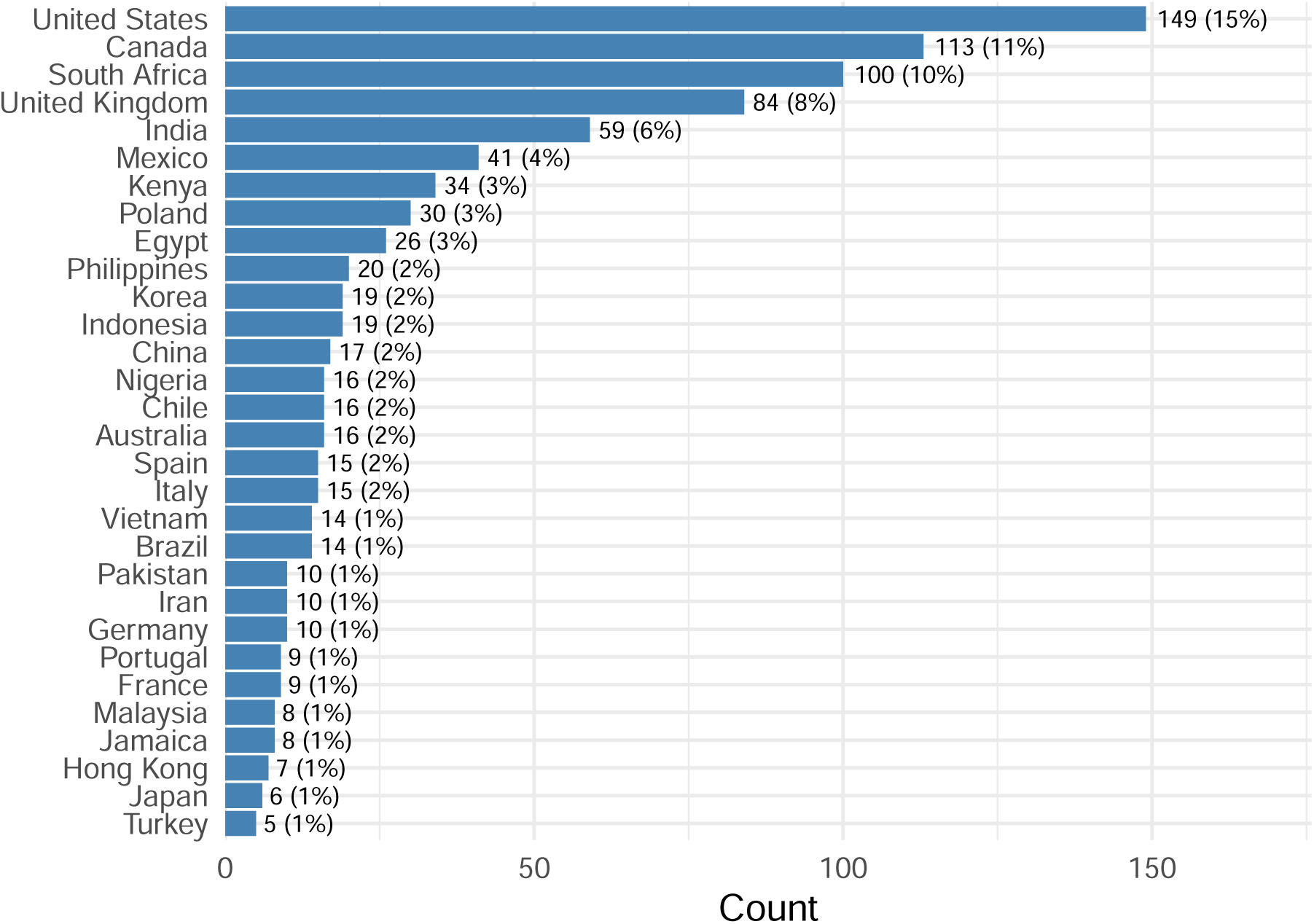
Respondents’ country of birth (top 30).

**Figure 6:**
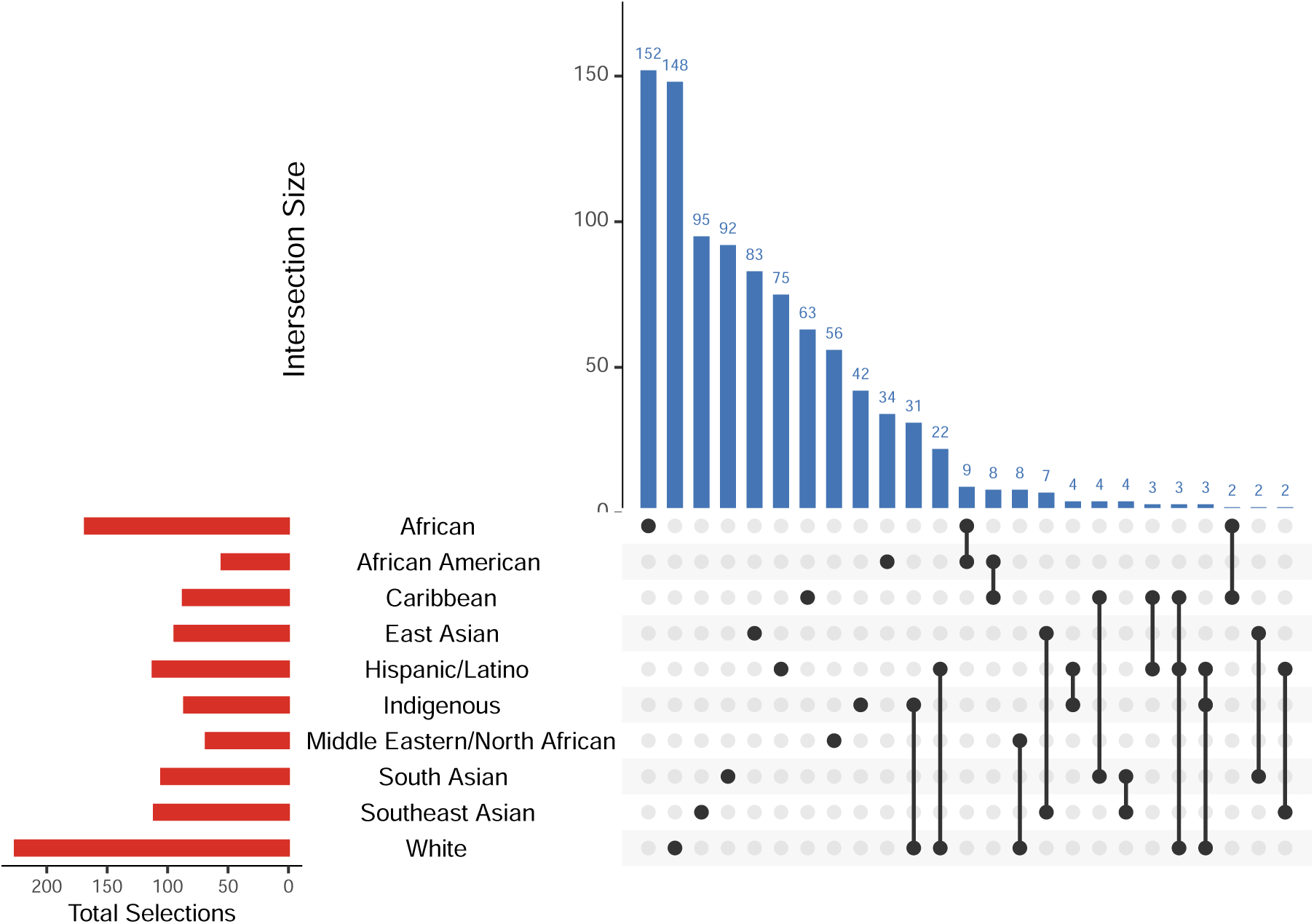
Intersection analysis of self-reported ethnic identities. Horizontal bars (left) show the total number of respondents selecting each ethnicity. Vertical bars (top) show the frequency of each unique combination of ethnicities, ordered by size; connected dots indicate multiethnic identifications.

**Figure 7:**
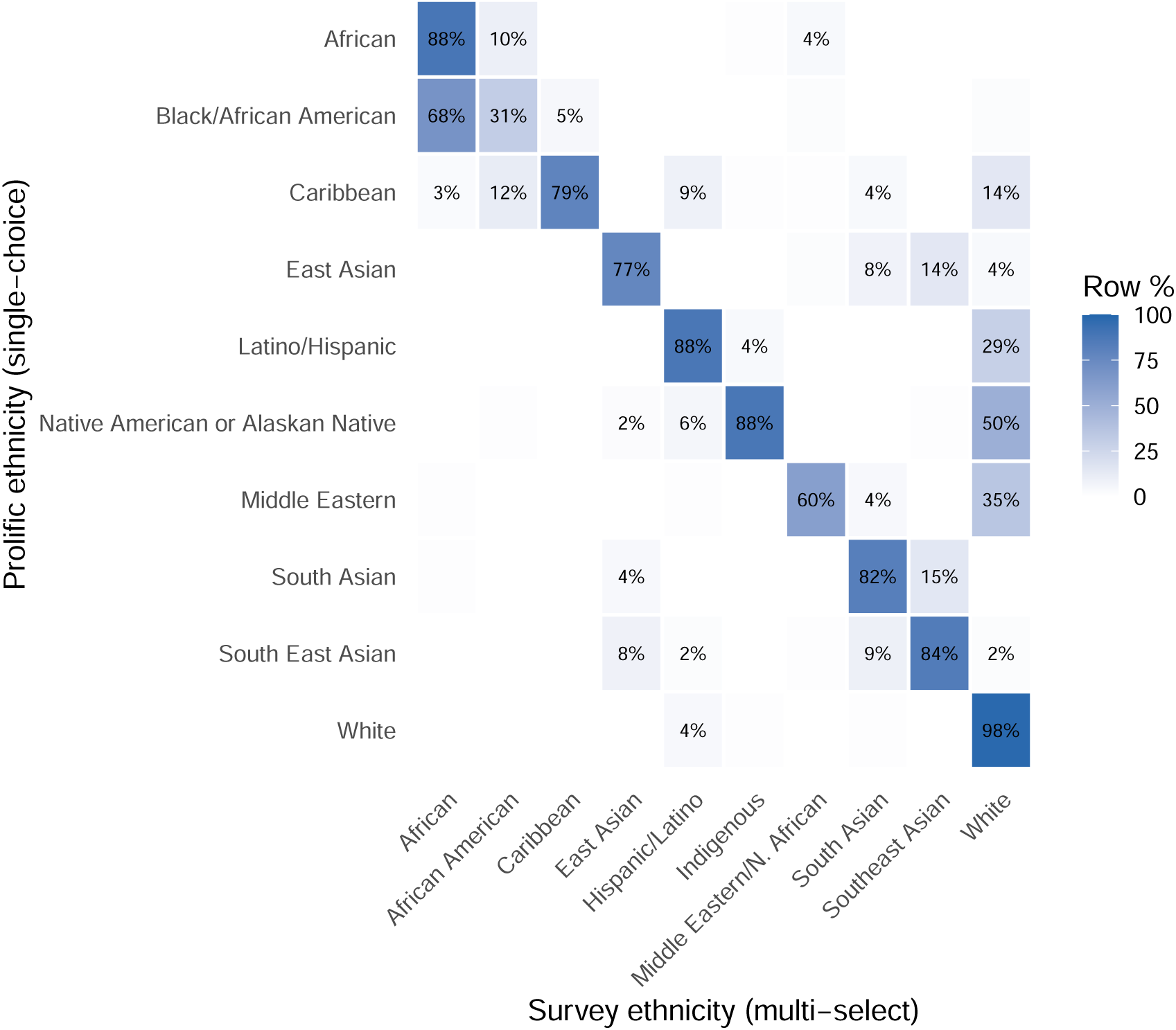
Participants’ self-reported ethnicity at platform sign-up (single-choice, rows) against their self-reported ethnicity in our survey (multi-select, columns). The sign-up classification was used for quota sampling. Cell colour and labels show the percentage of respondents in each sign-up category who selected a given survey ethnicity; row percentages can exceed 100% because survey responses were multi-select. Diagonal cells represent within-category concordance; off-diagonal cells reveal cross-category identification.

## Appendix D. Results Among Participants Who Experienced Racism

Table 5 compares the characteristics of participants who did and did not report unfair treatment in healthcare because of their race or ethnicity. Figure 8 shows their comfort with each calculator factor, Figure 9 their reactions to the race-specific algorithms, and Figure 10 their preferences among the revised algorithms.

**Table 5:**
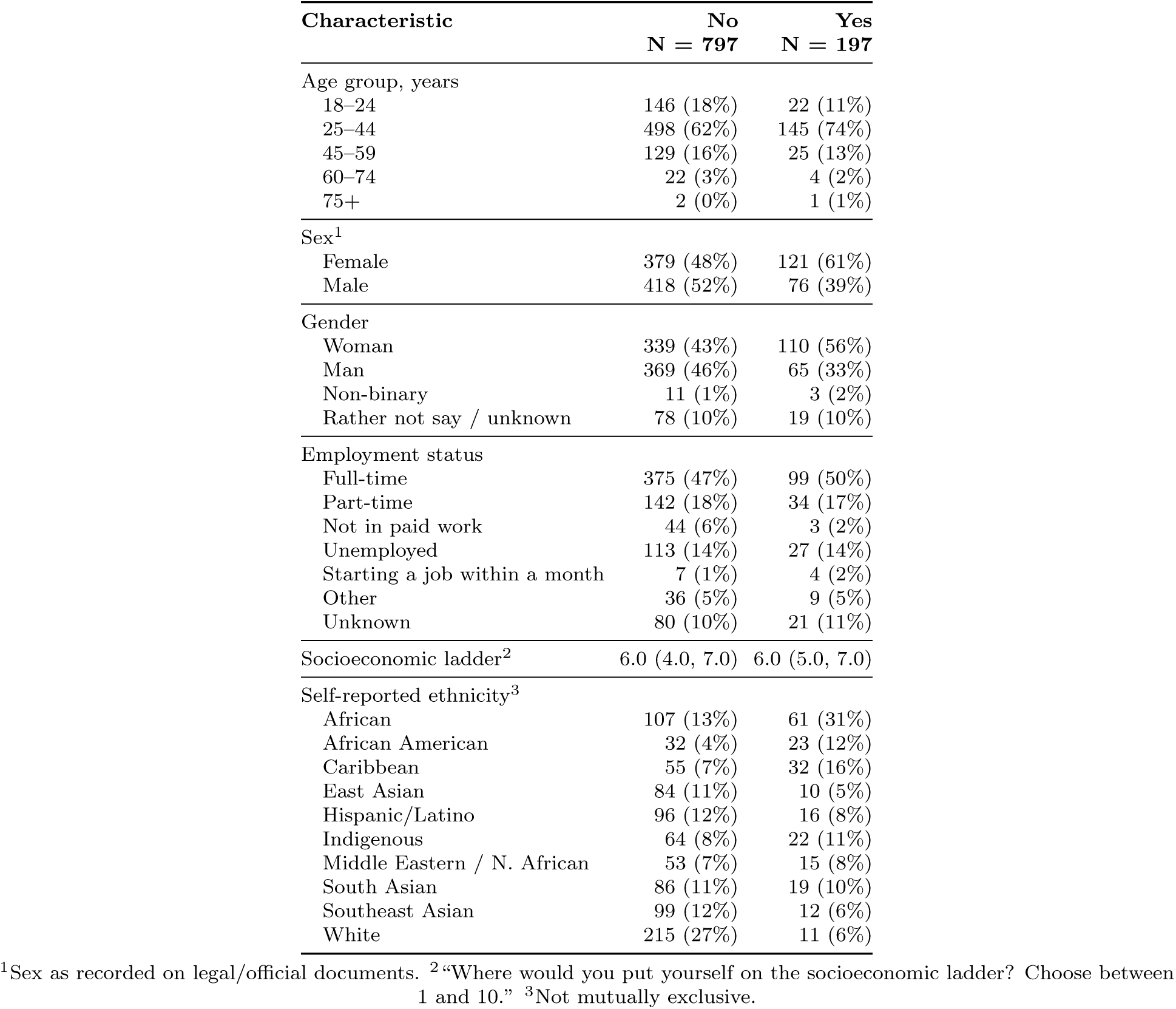
Characteristics of survey participants by whether they reported ever being treated unfairly in healthcare because of their race or ethnicity (”No” includes participants who answered Unsure or Prefer not to say). Ethnicity rows reflect the self-reported multi-select survey item; categories are not mutually exclusive, as respondents could select multiple identities. Values are n (%) or median (Q1, Q3).

**Figure 8:**
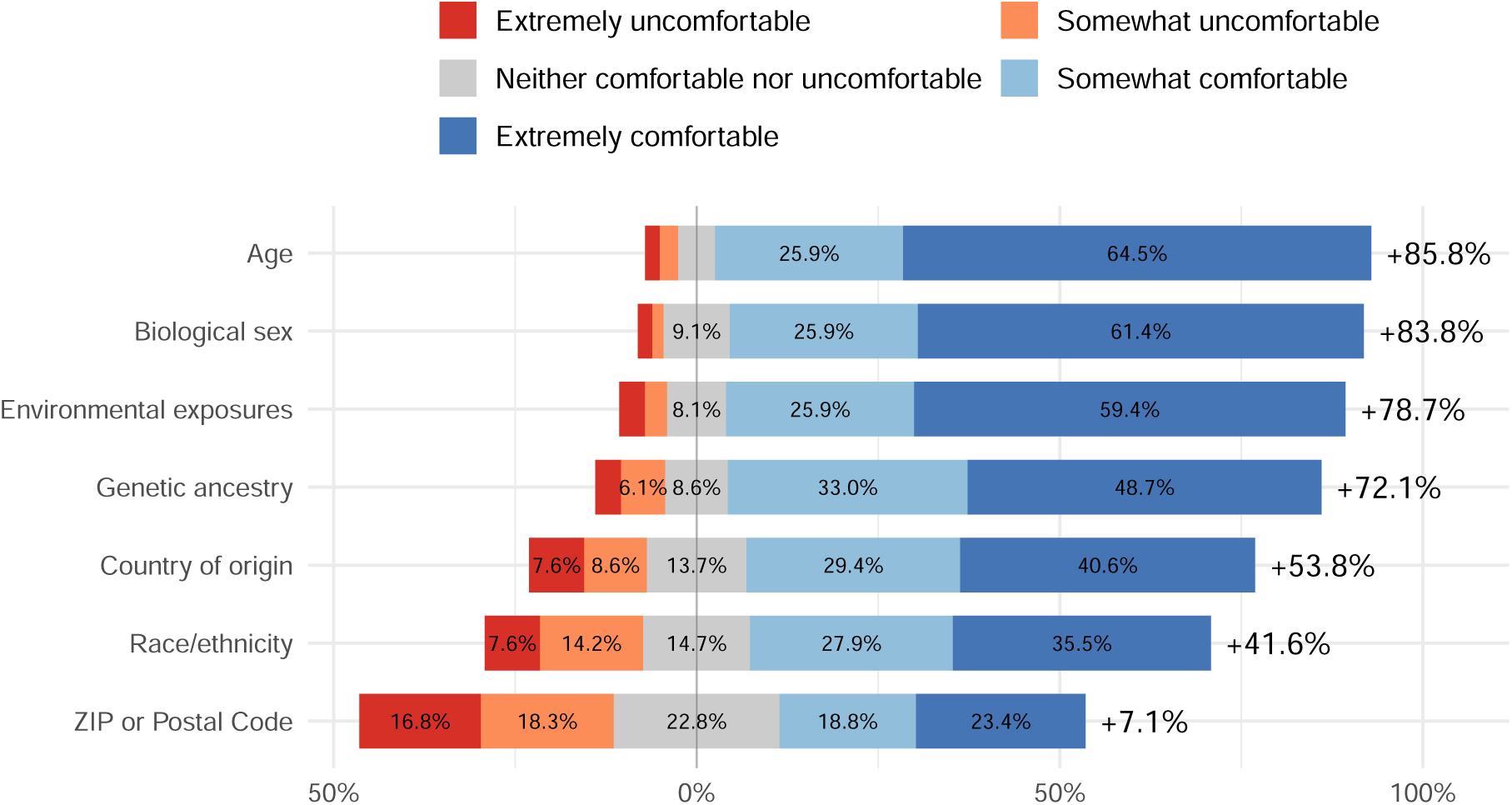
Comfort with calculator factors among respondents who reported unfair treatment due to race or ethnicity.

**Figure 9:**
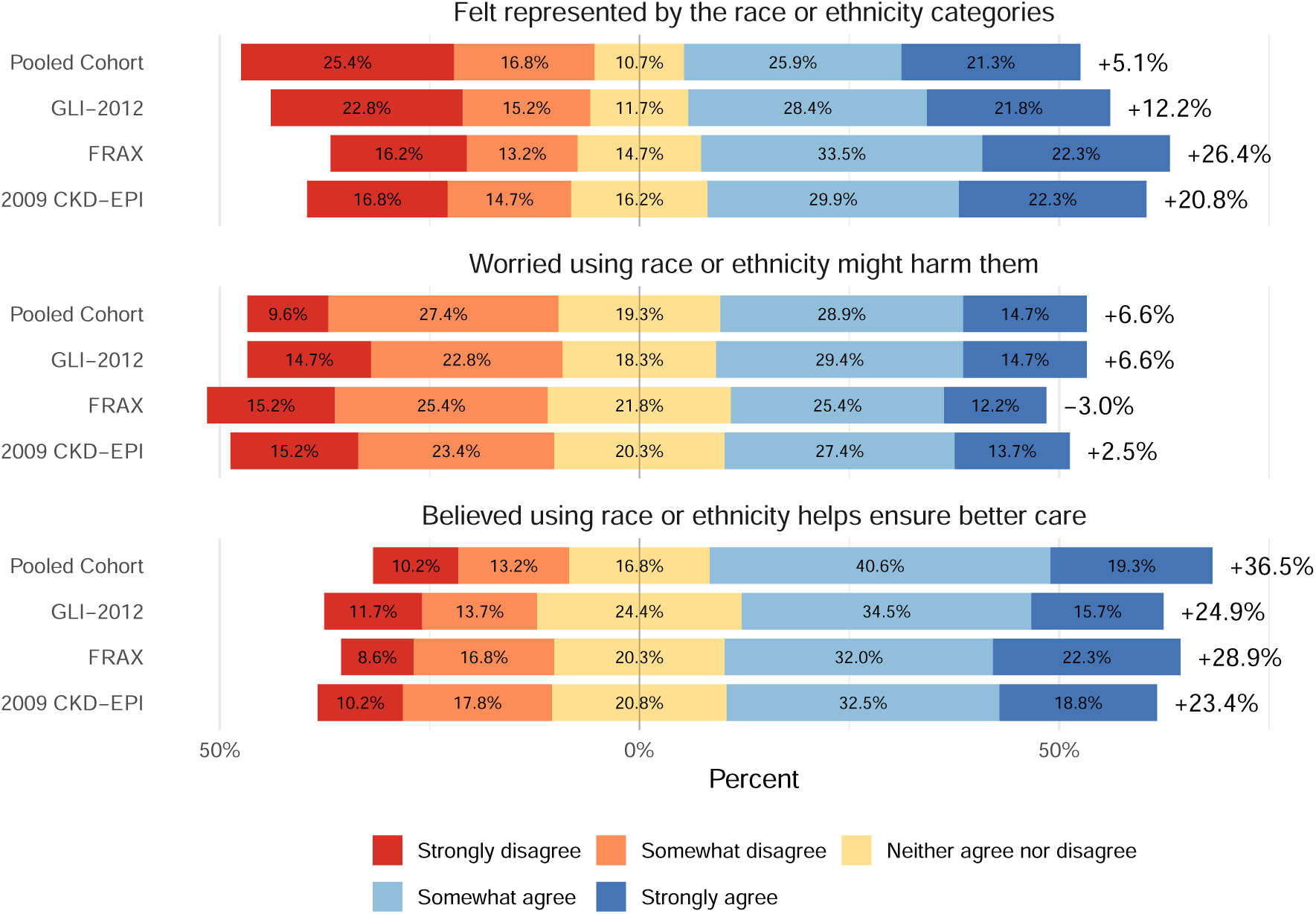
Reactions to race-specific algorithms among those who reported unfair treatment due to race or ethnicity. Bars extend rightward for agreement (light and dark blue) and leftward for disagreement (light and dark red), with neutral responses (yellow) centred at zero. Net agreement is shown to the right of each bar.

**Figure 10:**
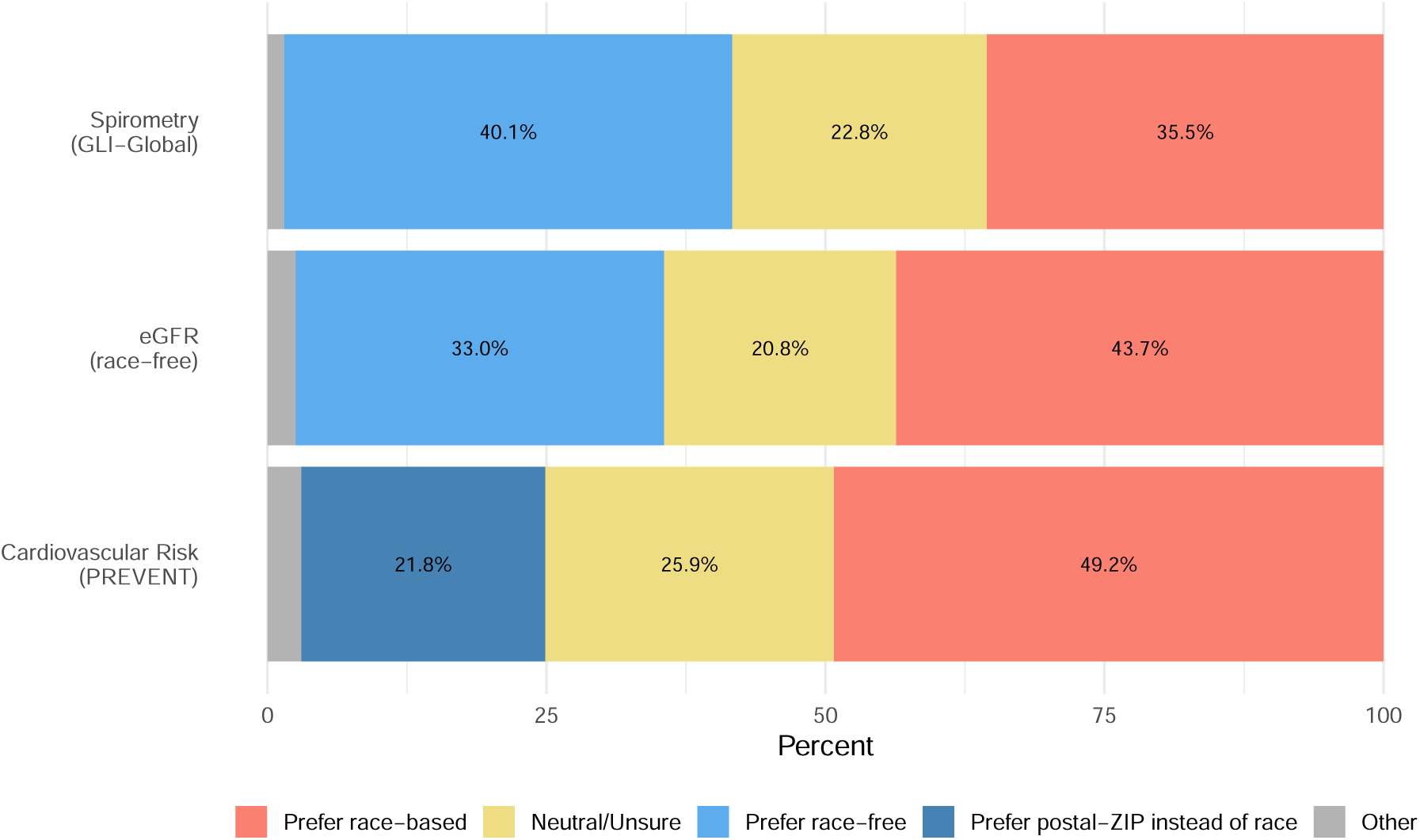
Reactions to the revised algorithms among those who reported unfair treatment due to race or ethnicity.

## Appendix E. Comfort by Ethnicity and Country

Figures 11 and 12 compare comfort with race or ethnicity, country of origin, and ZIP or postal code across self-reported and platform-recorded ethnicity groups; Figure 14 compares them across countries of residence. Figure 13 shows agreement, by ethnicity, with the statement that doctors should never use race or ethnicity in clinical algorithms.

**Figure 11:**
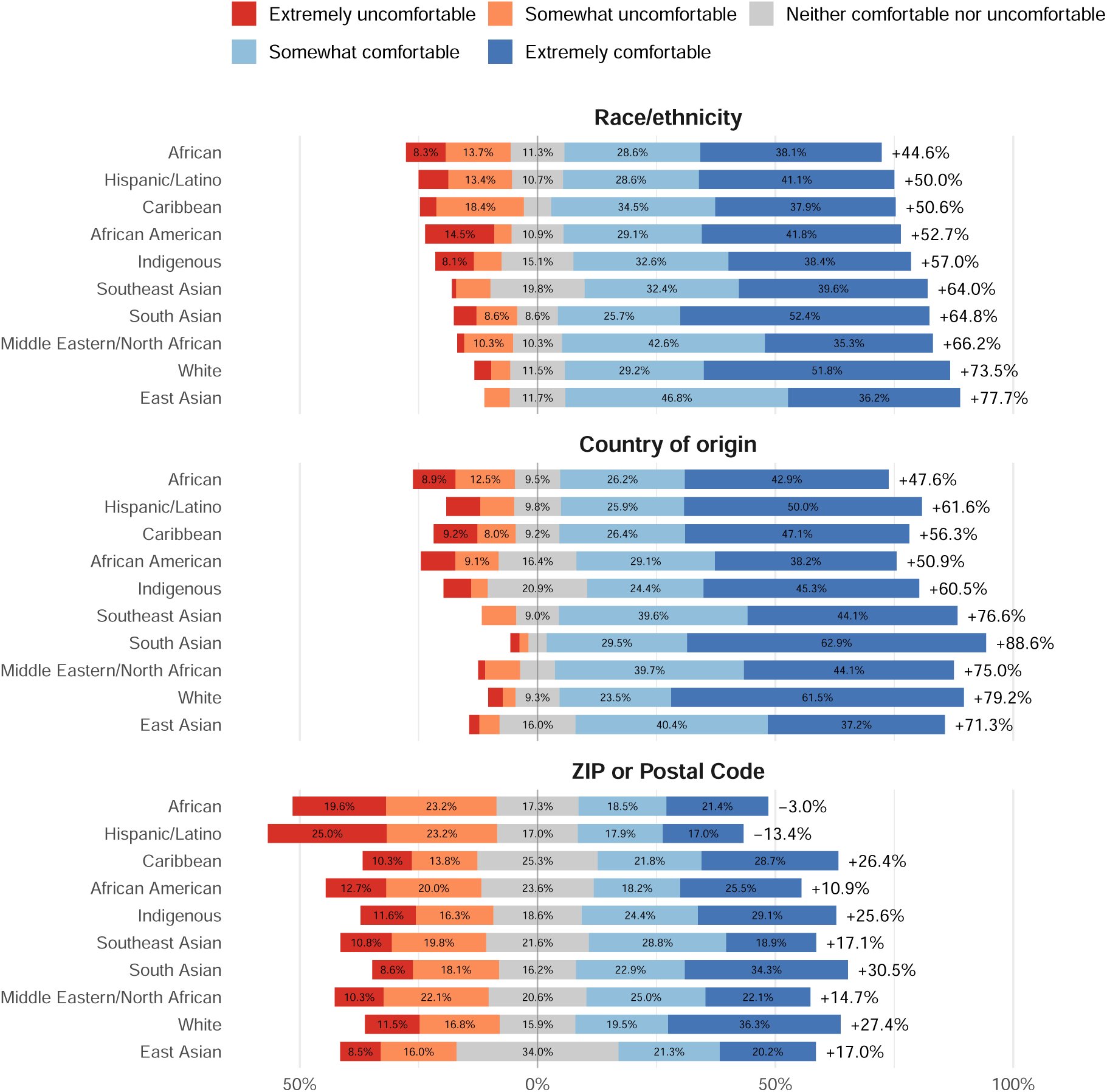
Comfort with race/ethnicity, country of origin, and ZIP or postal code as calculator factors, across self-reported ethnicity groups (multiple selections allowed). Respondents who selected multiple ethnicities are counted in each group they identified with. Groups are ordered from least to most comfortable with race/ethnicity. Bars extend rightward for comfortable and leftward for uncomfortable responses, with neutral responses centred at zero. Net comfort is shown to the right of each bar.

**Figure 12:**
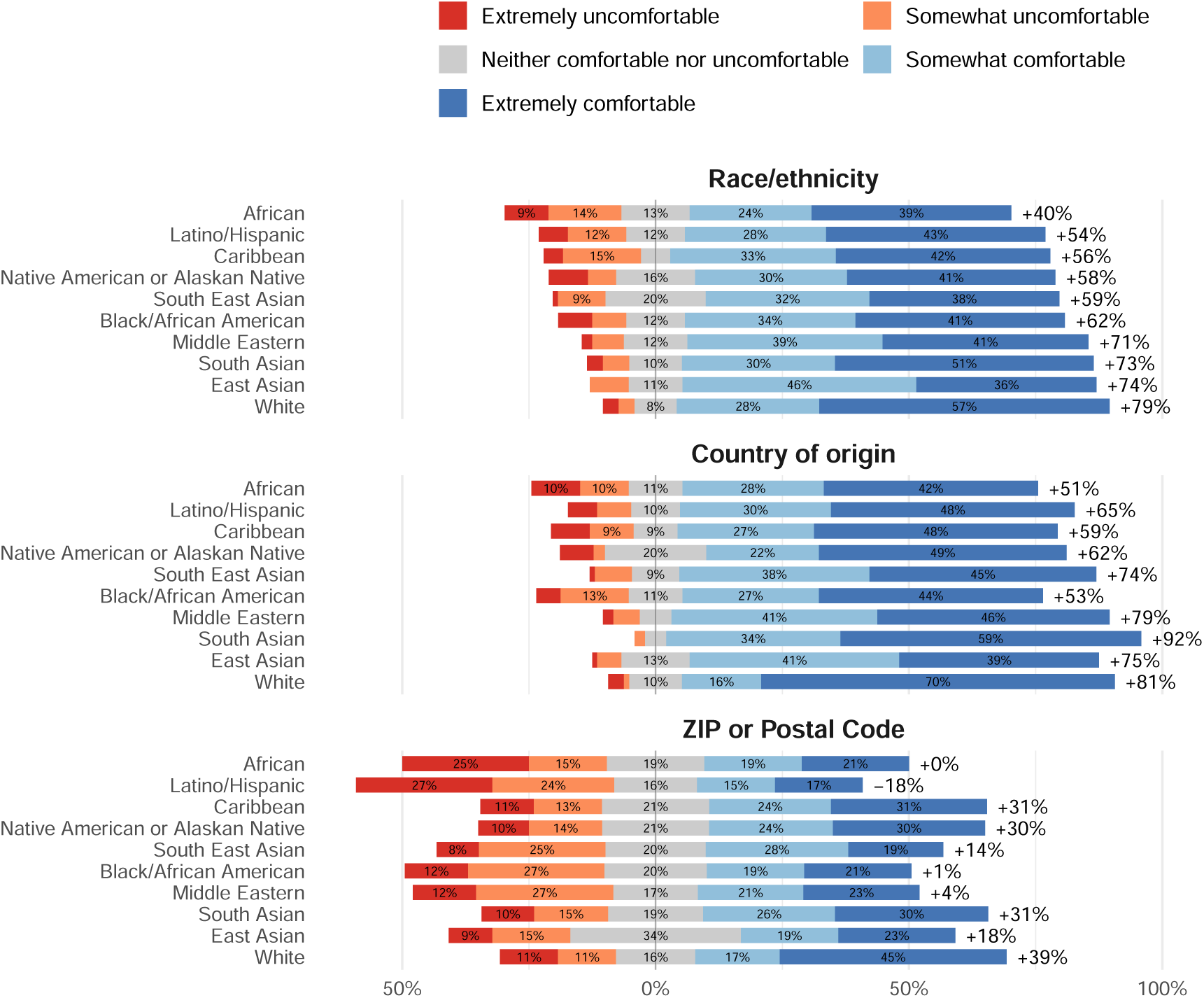
Comfort with race/ethnicity, country of origin, and ZIP or postal code as calculator factors, across ethnicity groups as recorded by the recruitment platform. Groups are ordered from least to most comfortable with race/ethnicity. Net comfort is shown to the right of each bar.

**Figure 13:**
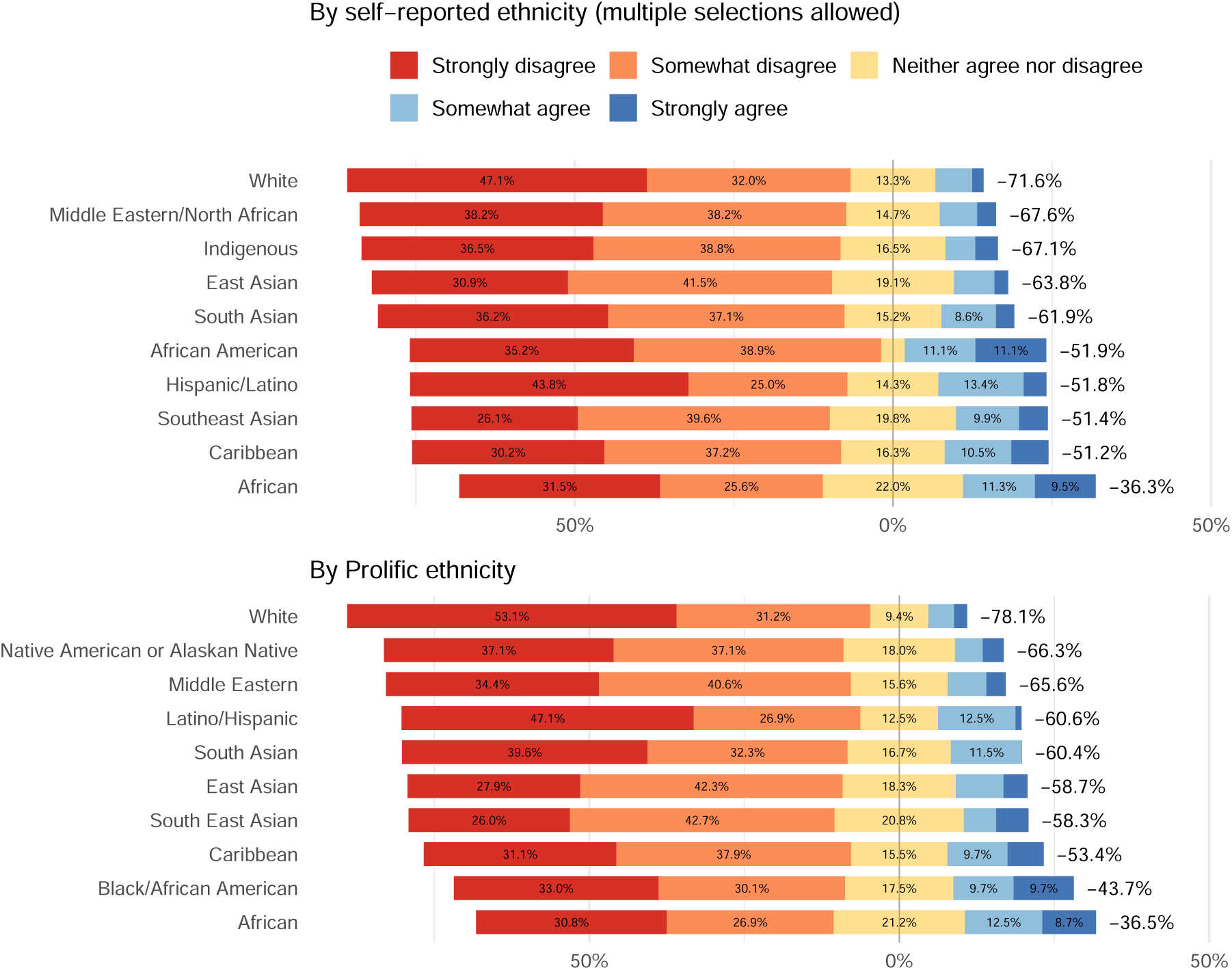
Agreement with the statement that doctors should never use race or ethnicity in clinical algorithms, by self-reported ethnicity (top, multiple selections allowed) and platform-recorded ethnicity (bottom). Bars extend rightward for agreement and leftward for disagreement, with neutral responses centred at zero. Net agreement is shown to the right of each bar.

**Figure 14:**
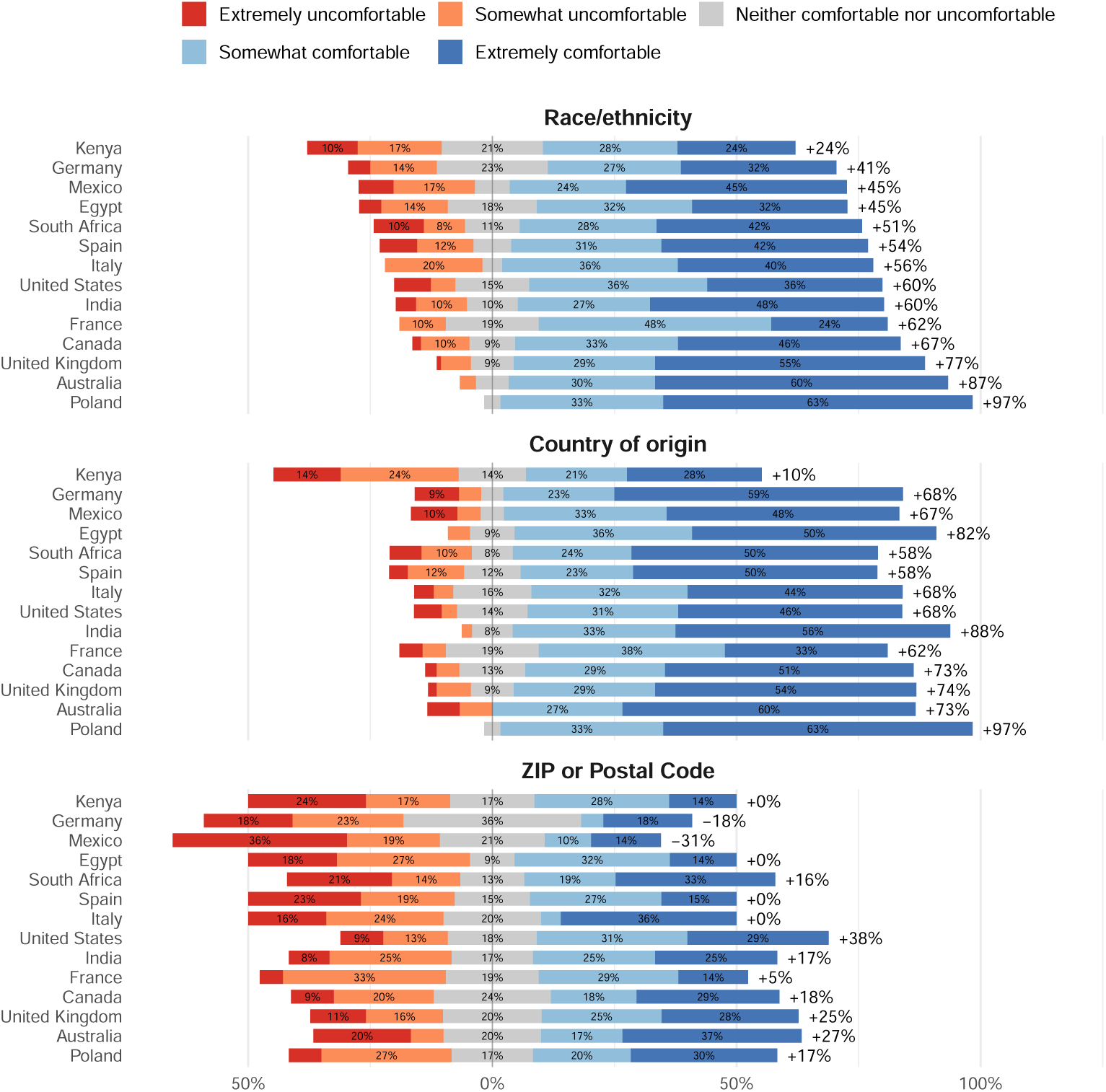
Comfort with race/ethnicity, country of origin, and ZIP or postal code as calculator factors, across countries of residence (countries with at least 20 respondents). Countries are ordered from least to most comfortable with race/ethnicity. Net comfort is shown to the right of each bar. Country estimates are descriptive and not nationally representative.

## Appendix F. Race and Ethnicity in Healthcare Encounters

Figures 15 and 16 show how often respondents are asked about their race or ethnicity when seeking healthcare, by ethnicity and by country. Figures 17 and 18 show how often clinic staff recorded race or ethnicity without asking, separating cases where the staff’s classification matched the respondent’s own choice from cases where it did not.

**Figure 15:**
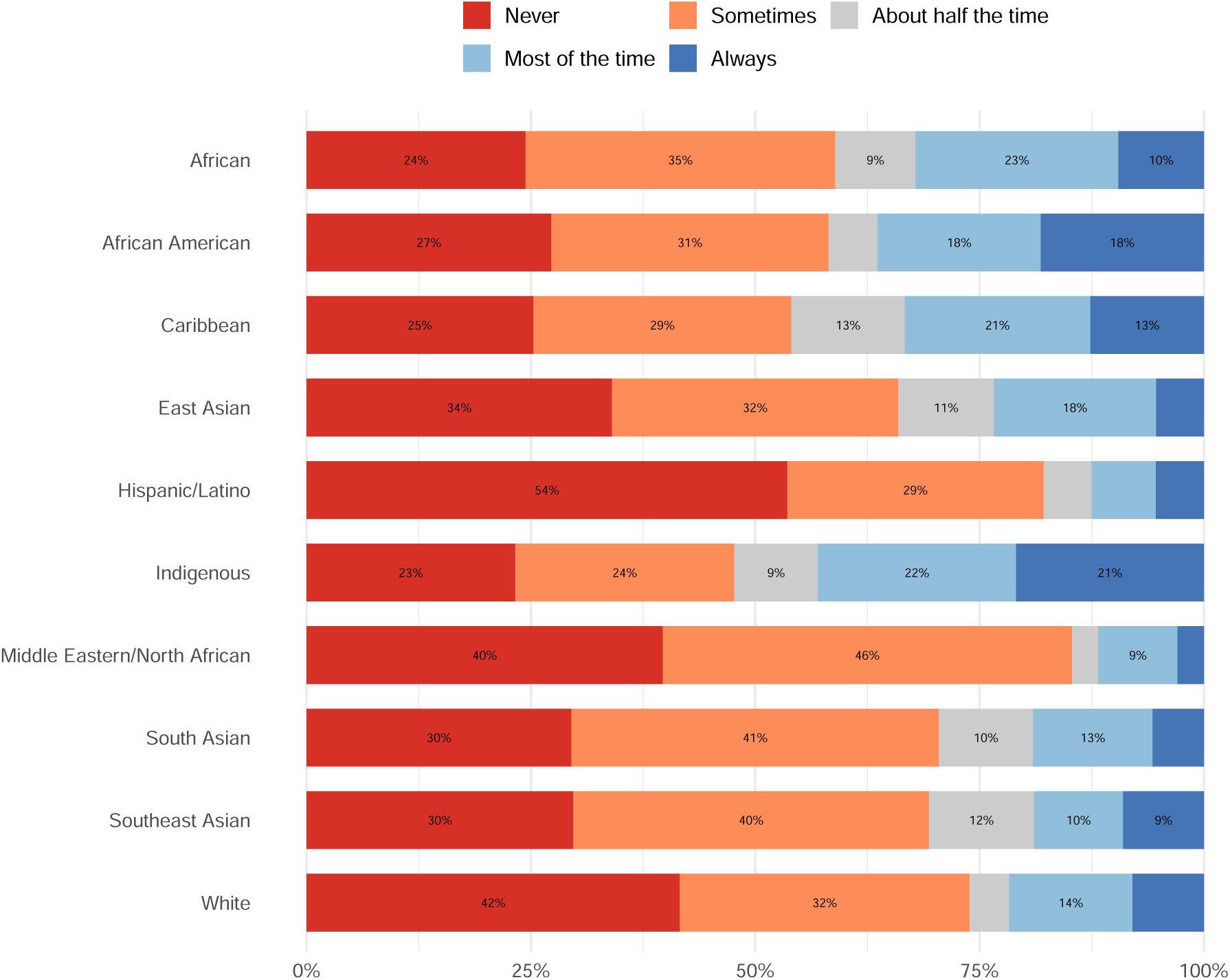
How often respondents are asked about their race or ethnicity when seeking healthcare, by self-reported ethnicity (multiple selections allowed). Respondents who selected multiple ethnicities are counted in each group they identified with. Groups are ordered alphabetically.

**Figure 16:**
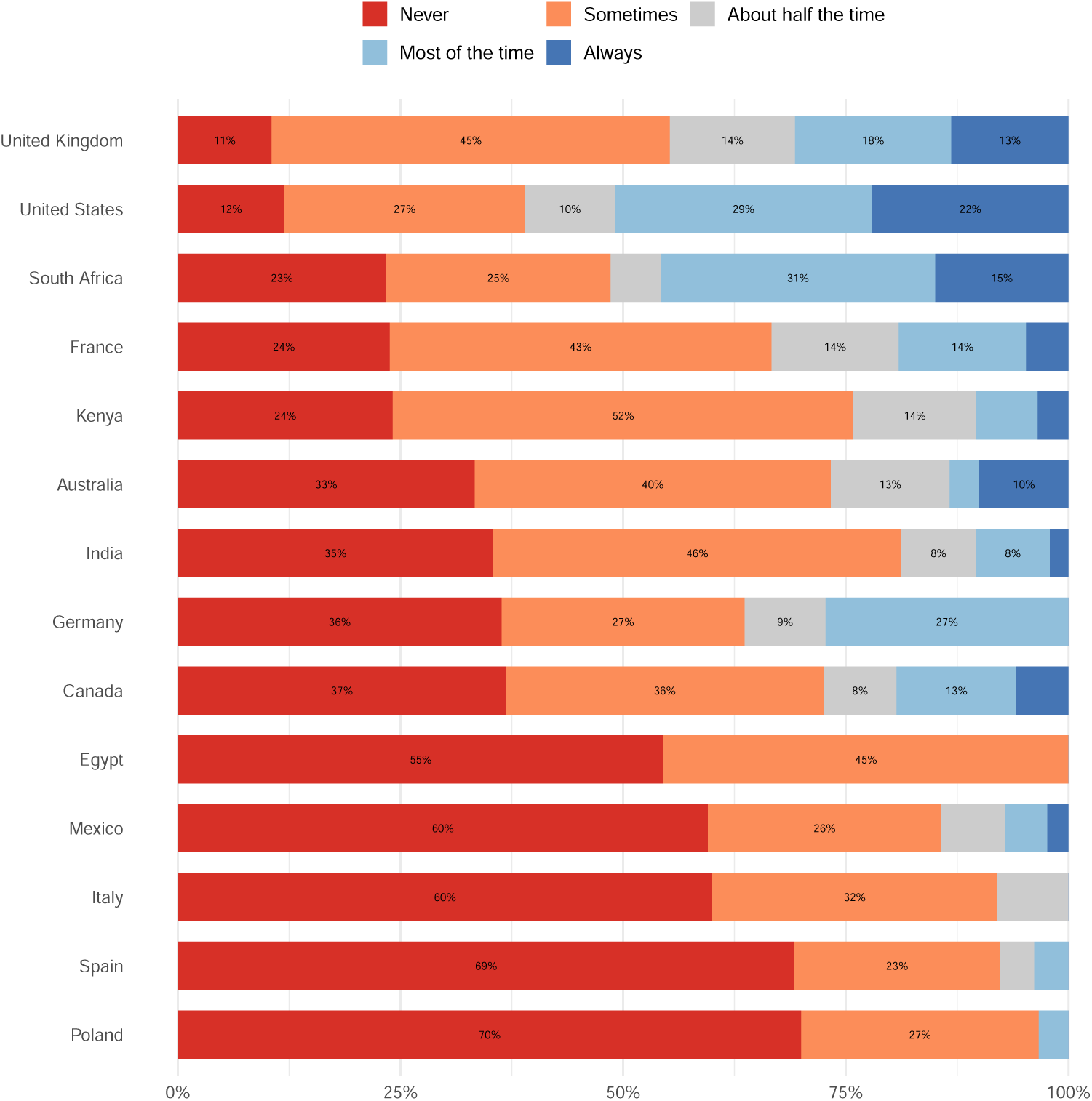
How often respondents are asked about their race or ethnicity when seeking healthcare, by country of residence (countries with at least 20 respondents). Countries are ordered from most to least frequently reporting “Never” being asked. Country estimates are descriptive and not nationally representative.

**Figure 17:**
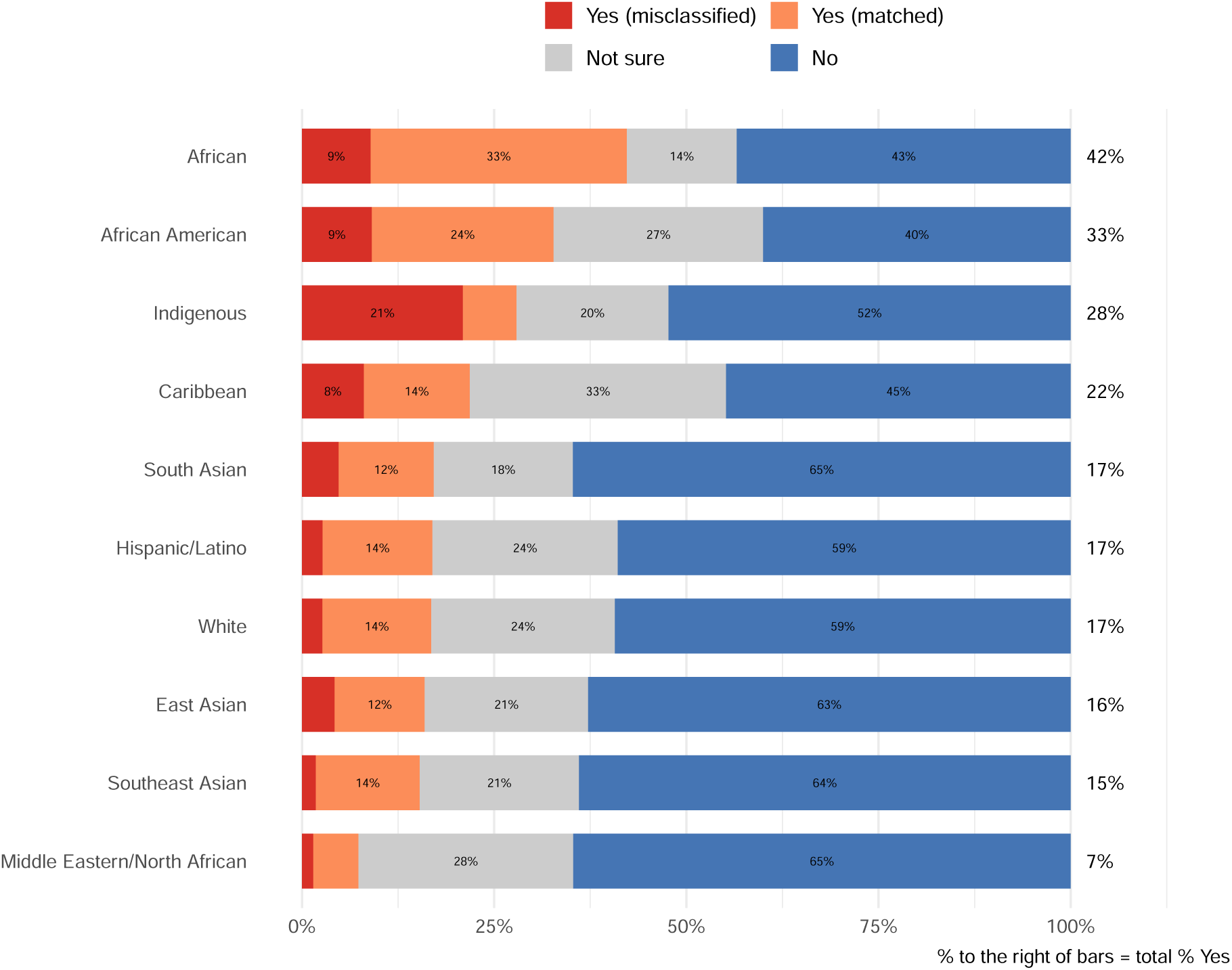
Proportion of respondents who reported that a doctor, nurse, or clinic staff member filled out their race or ethnicity without asking them, by self-reported ethnicity (multiple selections allowed). “Yes (matched)” indicates the staff classification matched the respondent’s own choice; “Yes (misclassified)” indicates it did not. Groups are ordered from lowest to highest total proportion answering “Yes”, shown to the right of each bar.

**Figure 18:**
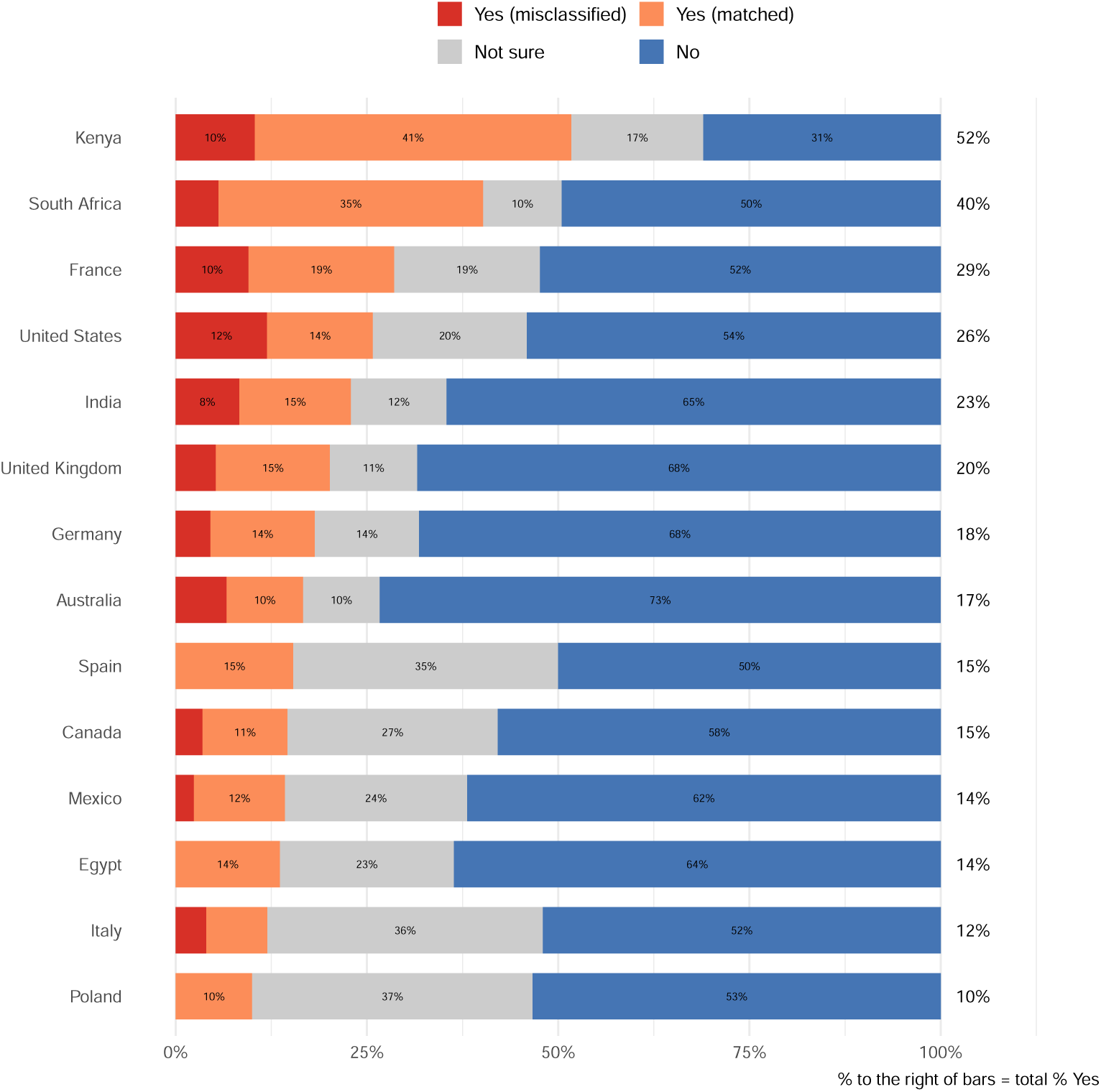
Proportion of respondents who reported that a doctor, nurse, or clinic staff member filled out their race or ethnicity without asking them, by country of residence (countries with at least 20 respondents). Countries are ordered from lowest to highest total proportion answering “Yes”, shown to the right of each bar. Country estimates are descriptive and not nationally representative.

## Appendix G. Comparison with Representative US Surveys

To gauge potential bias in our multinational convenience sample, we replicated questions from representative US surveys. The item “In health and medicine, how much of a problem is bias and unfair treatment based on patients’ race or ethnicity?” was adapted from the Pew Research Center (Tyson et al., 2023), and “In general, do you think you would receive better or worse care from doctors or health care providers who share your racial and ethnic background, or would it not make much difference?” from the Kaiser Family Foundation (Kaiser Family Foundation, 2026). Figures 19 and 20 compare our overall and US-resident responses with those representative samples.

We also replicated items from Diao et al. (2026), who surveyed a representative US sample on comfort with demographic factors in a cancer risk calculator and on attitudes toward the use of race in clinical care. Figure 21 compares comfort with the overlapping factors (age, sex, race, and ZIP or postal code), and Figure 22 compares agreement with four statements about using race in clinical care.

**Figure 19:**
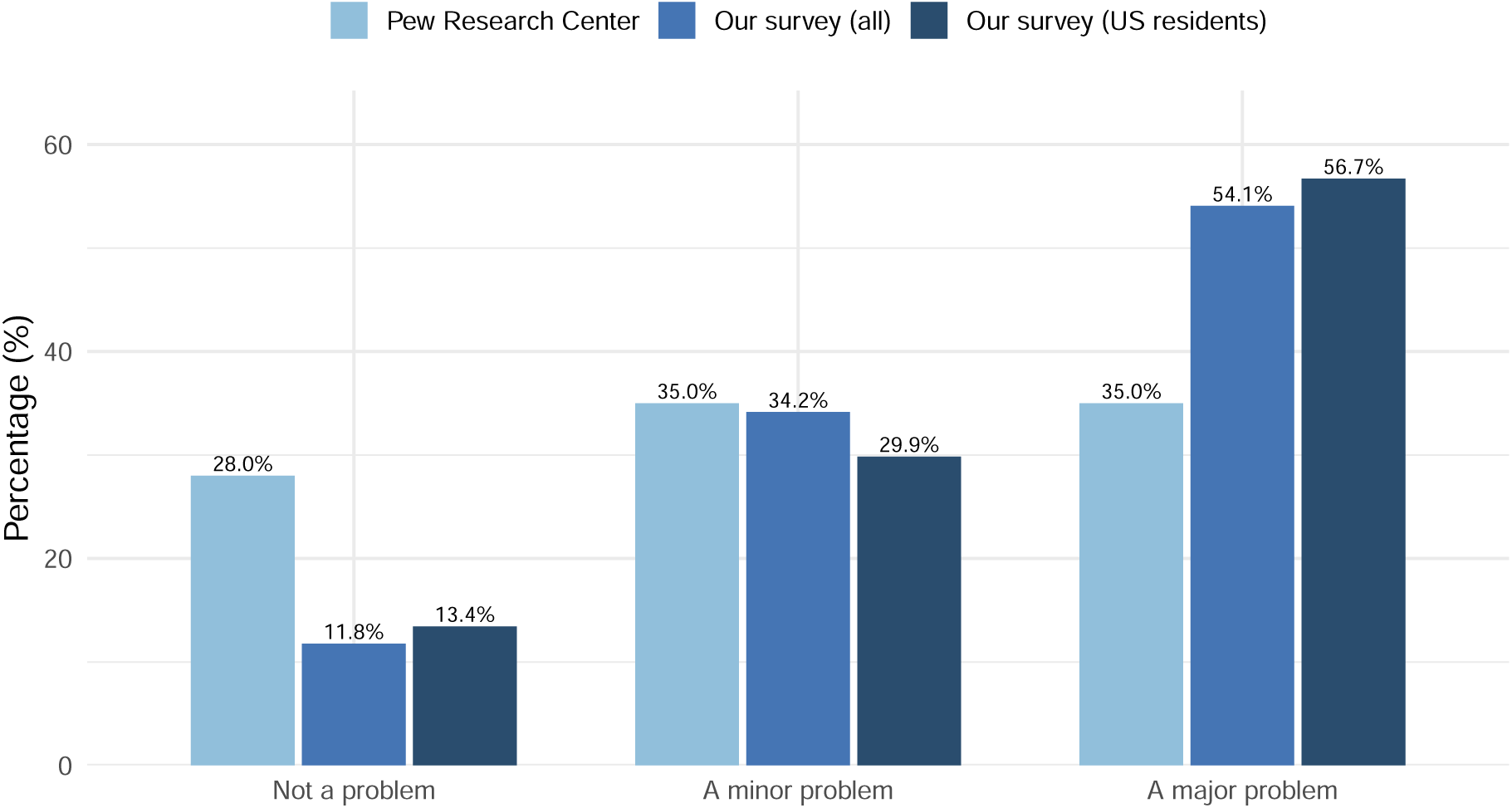
Responses to “In health and medicine, how much of a problem is bias and unfair treatment based on patients’ race or ethnicity?” in our survey (all respondents and US residents only) versus a representative US sample from the Pew Research Center. Respondents answering “Unsure” in our survey were excluded, as that option was not offered in the Pew survey.

**Figure 20:**
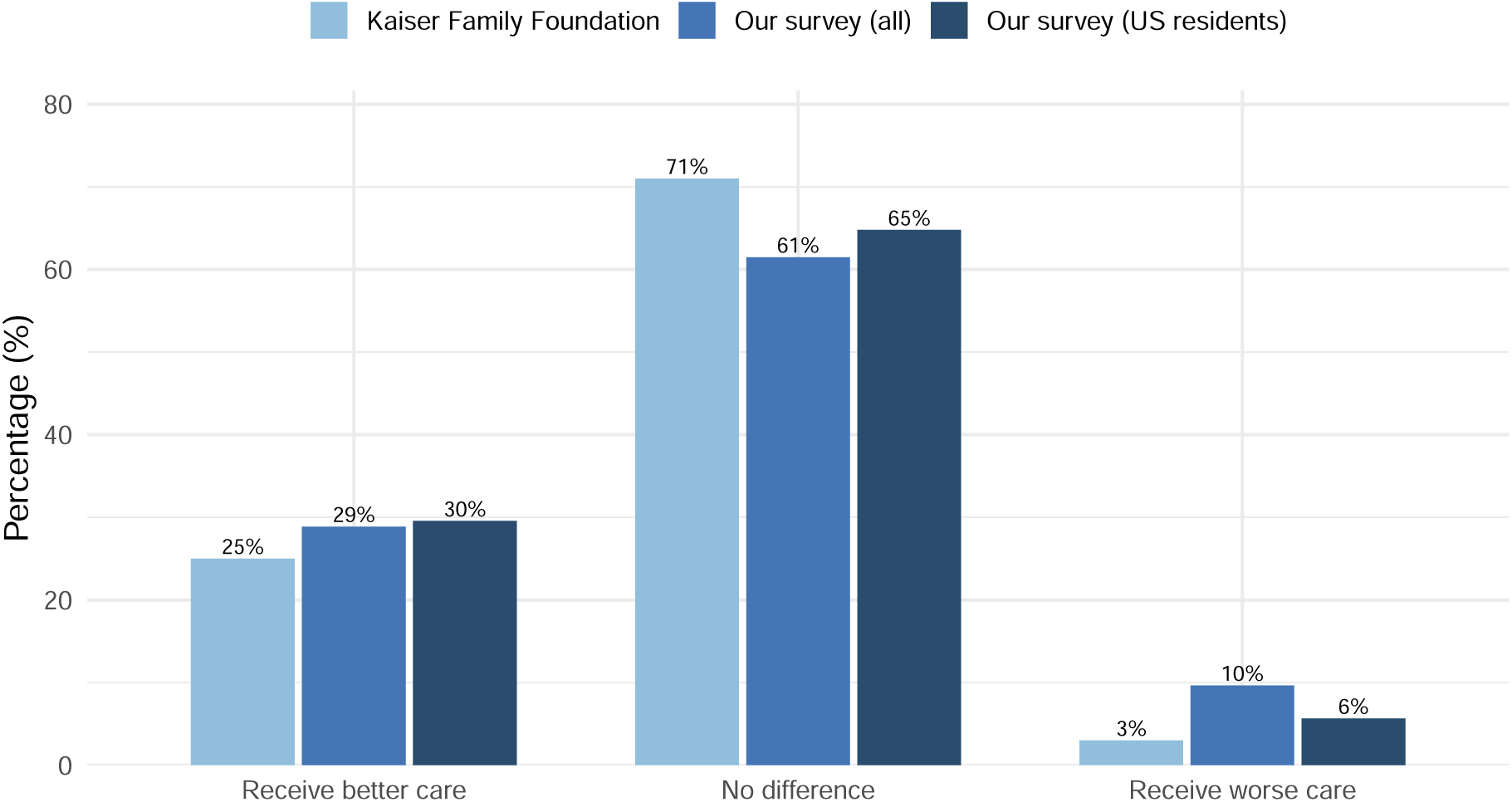
Responses to “Do you think you would receive better or worse care from doctors or health care providers who share your racial and ethnic background, or would it not make much difference?” in our survey (all respondents and US residents only) versus a representative US sample from the Kaiser Family Foundation.

**Figure 21:**
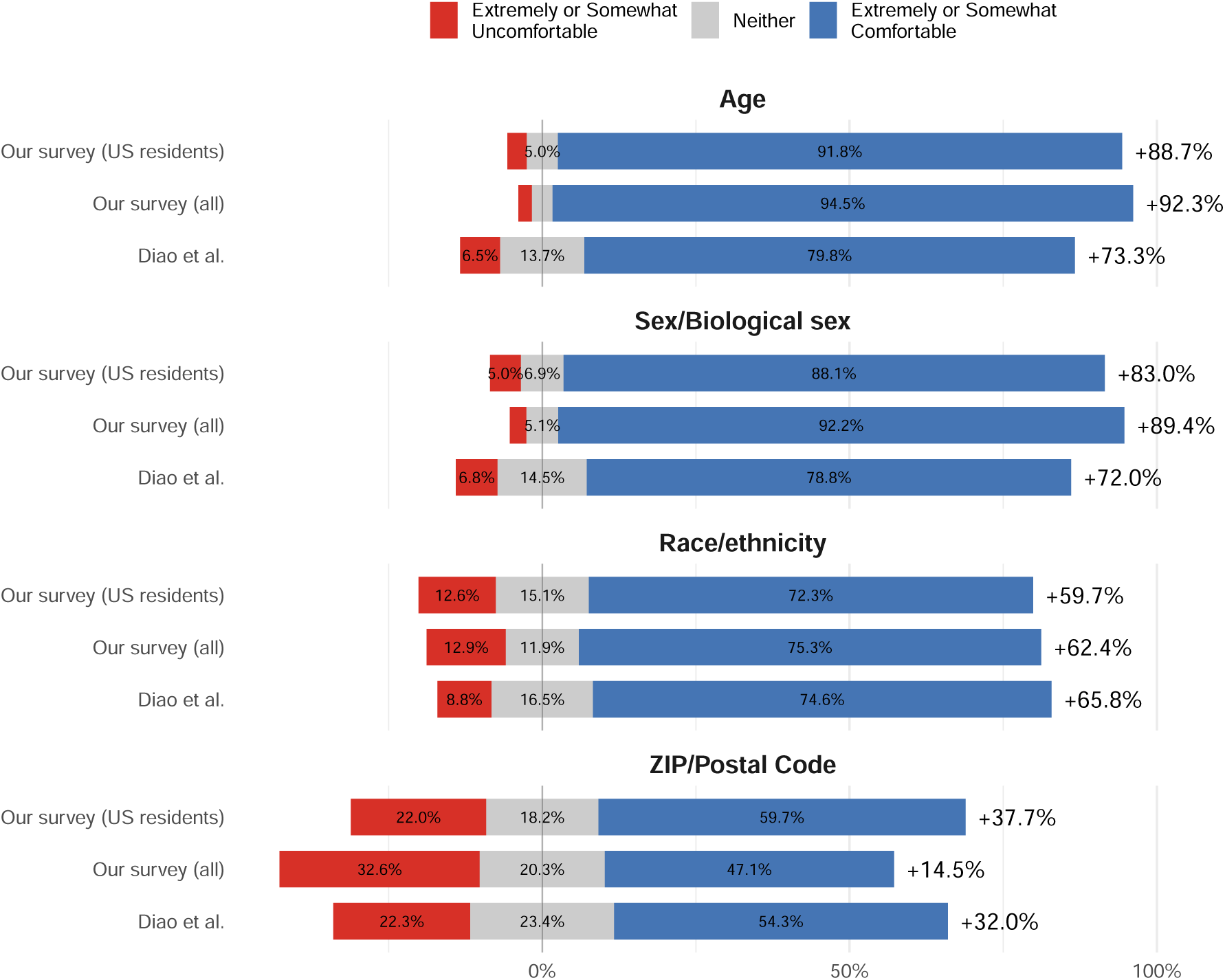
Comfort with demographic factors in a hypothetical risk calculator in our survey (all respondents and US residents only) versus the representative US sample of Diao et al. (2026). Bars extend rightward for comfortable (blue) and leftward for uncomfortable (red) responses, with neutral responses (grey) centred at zero. Net comfort is shown to the right of each bar. Responses were collapsed into three categories to match the reporting in that study.

**Figure 22:**
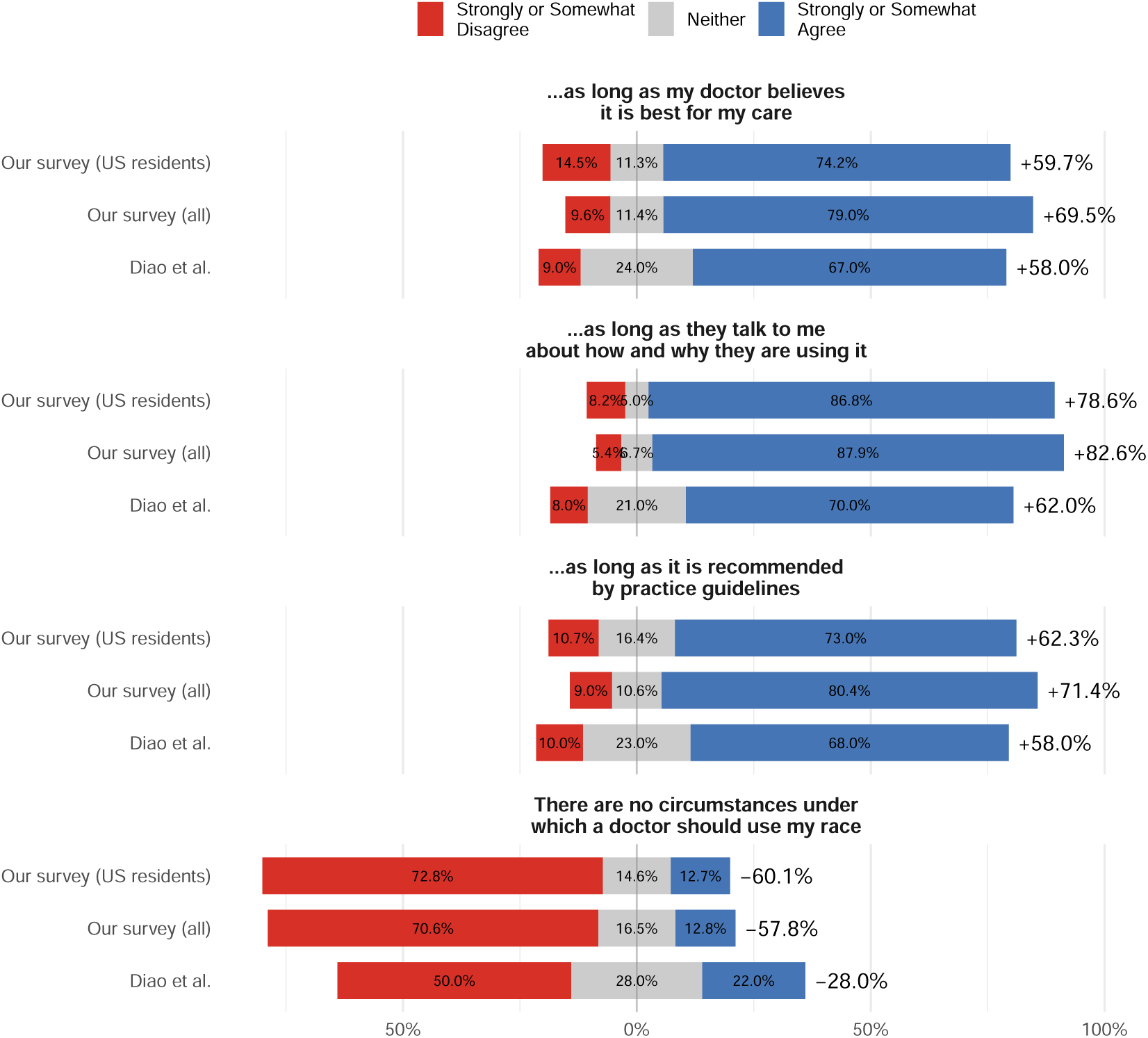
Agreement with statements about the use of race or ethnicity in clinical care in our survey (all respondents and US residents only) versus the representative US sample of Diao et al. (2026). The first three statements are prefixed with “I am comfortable with my doctor using my race or ethnicity in clinical care. . . ” Net agreement is shown to the right of each bar. Responses were collapsed into three categories to match the reporting in that study.

## Appendix H. Qualitative Code Frequencies and Inter-rater Agreement

Table 8 reports the frequency of each code across the three open-ended questions. Codes are not mutually exclusive—a single response could receive several—so percentages do not sum to 100%. Additional example quotes for each question are provided in Appendix I.

Two reviewers independently coded 10% of responses to each question (100, 10, and 23 responses). Table 6 reports pooled inter-rater agreement over all response-by-code decisions, and Table 7 reports agreement for each code. Because *κ* is unstable for rarely applied codes—it is 0 when one reviewer applies a code once and the other never, and undefined when neither applies it—we report Gwet’s AC1 alongside it. Per-code intervals are wide for rare codes, particularly for accounts of racism (10 responses), and should be read with caution. All discrepancies were resolved by consensus before the remaining responses were coded by one reviewer.

**Table 6:** Pooled inter-rater agreement across all response-by-code decisions, with 95% percentile bootstrap confidence intervals (2,000 resamples of responses).

| **Question** | **Agreement, %** | **Cohen's *κ*** | **Gwet's AC1** |
|---|---|---|---|
| Meaning of race and ethnicity | 94.6 | 0.79 | 0.93 |
|  | (93.1–96.1) | (0.74–0.84) | (0.91–0.95) |
| Accounts of racism | 92.9 | 0.77 | 0.90 |
|  | (88.6–97.1) | (0.64–0.91) | (0.83–0.96) |
| Concerns about reform | 96.2 | 0.71 | 0.96 |
|  | (94.2–98.0) | (0.55–0.85) | (0.93–0.98) |
| All questions | 94.9 | 0.79 | 0.93 |
|  | (93.7–96.1) | (0.74–0.83) | (0.92–0.95) |

**Table 7:**
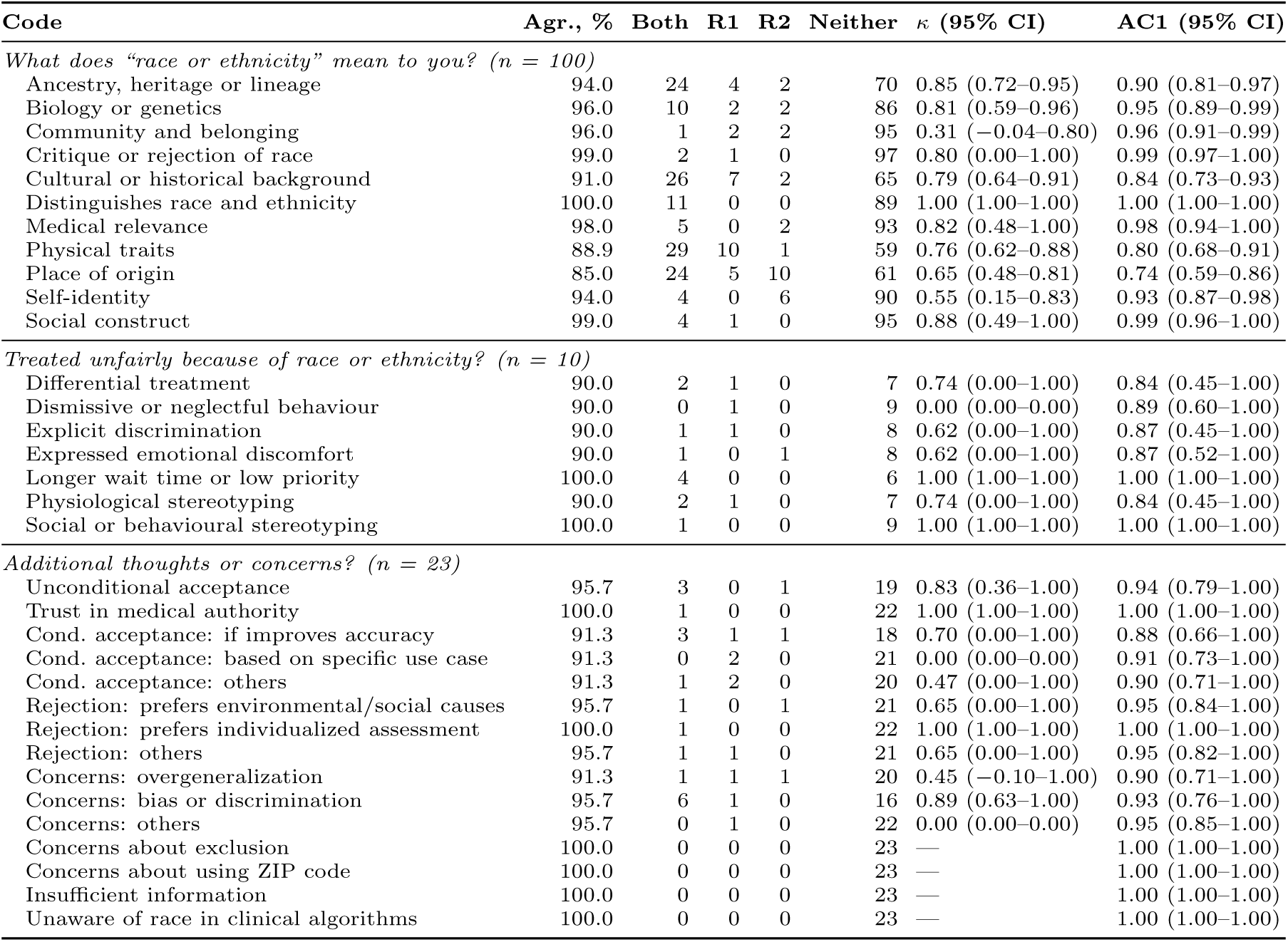
Inter-rater agreement for each code, with 95% percentile bootstrap confidence intervals (2,000 resamples of responses). Both: responses coded by both reviewers; R1, R2: responses coded by reviewer 1 or 2 only; Agr.: percent agreement; *κ*: Cohen’s kappa; AC1: Gwet’s AC1. —: *κ* undefined because neither reviewer applied the code. One missing rating (Physical traits) was excluded.

**Table 8:** Frequency of codes and categories identified across open-ended responses. Percentages are the proportion of total responses analyzed for that question.

| **Category** | **n** | **%** |
|---|---|---|
| *What does “race or ethnicity” mean to you? (n = 993)* |  |  |
| Physical traits | 370 | 37.3 |
| Cultural or historical background | 293 | 29.5 |
| Place of origin | 266 | 26.8 |
| Ancestry, heritage or lineage | 226 | 22.8 |
| Biology or genetics | 169 | 17.0 |
| Distinguishes race and ethnicity | 123 | 12.4 |
| Community and belonging | 70 | 7.0 |
| Critique or rejection of race | 53 | 5.3 |
| Medical relevance | 45 | 4.5 |
| Self-identity | 37 | 3.7 |
| Social construct | 29 | 2.9 |
| *Treated unfairly because of race or ethnicity? (n = 90)* |  |  |
| Differential treatment | 27 | 30.0 |
| Dismissive or neglectful behaviour | 26 | 28.9 |
| Longer wait time or low priority | 20 | 22.2 |
| Physiological assumption or stereotyping | 15 | 16.7 |
| Social or behavioural stereotyping | 7 | 7.8 |
| Explicit discrimination | 6 | 6.7 |
| Expressed emotional discomfort | 5 | 5.6 |
| *Additional thoughts or concerns? (n = 215)* |  |  |
| Unconditional acceptance | 37 | 17.2 |
| Trust in medical authority | 6 | 2.8 |
| *Conditional acceptance* |  |  |
| If improves accuracy | 19 | 8.8 |
| Based on specific use case | 17 | 7.9 |
| Others | 13 | 6.0 |
| *Rejection* |  |  |
| Prefers environmental/social causes | 13 | 6.0 |
| Prefers individualized assessment | 13 | 6.0 |
| Others | 14 | 6.5 |
| *Concerns about using race* |  |  |
| Overgeneralization | 15 | 7.0 |
| Bias or discrimination | 24 | 11.2 |
| Others | 12 | 5.6 |
| Concerns about exclusion | 14 | 6.5 |
| Concerns about using ZIP code | 2 | 0.9 |
| Insufficient information to form an opinion | 16 | 7.4 |
| Unaware of race in clinical algorithms | 10 | 4.7 |

## Appendix I. Additional Example Quotes

### Defining Race or Ethnicity

The excerpts below are drawn from participant responses to the following open-ended question: “In your own words, what does ‘race or ethnicity’ mean to you? There are no right or wrong answers - we are interested in your personal perspective.”

“For me, race is my physical traits, how do I look. Ethnicity is my heritage, how do I feel. For example, I am Southeast Asian with light brown skin, black hair, light brown eyes. My mom’s ethnicity is Batak Simalungun, while my dad’s is Batak Tapanuli. I was raised in Simalungun and spent half of my life here. So, I considered my ethnicity as a Simalungunese, although my surname is from Tapanuli.”

“Ancestors/cultural background - I’m Inuit, and native people in general are excluded too often in medical settings/research/general consideration. Indigenous people are so important to medical research and care, and it’s unethical to exclude us. Many institutions can and should do better (including you guys, perhaps, since you did not specifically include us); we should not be lumped into a general ‘other’ group, and as the original peoples of North America, it’s wrong on so many levels for research, etc to not consider our medical backgrounds and needs. I think it’s very important to consider Black, Asian, etc health systems, family history, etc as well, but please stop excluding the natives!”

“Race or ethnicity means a whole lot to me. It is one of those factors that signal proximity to a person’s identity. Race forms a huge part in my identity. For example, i am a black african male before i am anything else, and this is incredibly important to me because it pours out that i have experiences that are reflective to black people rather than any other race.”

“To me, race or ethnicity means clanship and togetherness, a feeling of belonging to people who share the same culture, traditions, and background. It reflects common roots and a shared identity that connects individuals as part of a community.”

“Race is a social construct used to make some feel superior to others. Ethnicity is where you were born or the cultures you grow up with. For example, if black people have a higher incidence of lung or kidney disease it could be because they live in red lined districts, with poor air and water quality regulations. It’s not because they’re black that their health is bad, it’s because health and safety measures and social constructs have placed them in this predicament.”

“In healthcare, race and ethnicity means ‘genetic background’.”

“Mostly just skin color and/or an area of origination of a certain people; but even then. . . they aren’t the same. Are North American indigenous the same as Australian aborigines, or even S. American indigenous people? What about Japanese vs Korean vs Chinese vs Vietnamese vs Filipino? They’re quite different, but some of these people are mixed blood as well. Tbh, race or ethnicity is just a generalization for an unknown, and just a guess about where someone may originate from.”

“Race or ethnicity feels like family to me, yet also like distant relatives. When I go back to my hometown, I feel no particular connection because everyone is the same race. But when I go to another country and see someone of my own race, I can’t tell you how glad I am to see them. That’s how race feels to me—like family, yet like distant relatives.”

“As an East Asian, I do really care and concern about my ethnicity and race, and prejudice/discrimination against my race since I move to an European country. Therefore, race or ethnicity is something threaten me and make me worried and scared in daily life. I do have rather negative image for it.”

“It is a huge part of who I am and dare I say one of the most important. I am German and Native American. The very dichotomy of that mixture has caused so much contradiction not only internally but externally as well. I never quite understood it until I got older. I always felt drawn to choose one. Looking too white to be native and looking too native to be white. I never understood why it mattered because I was still me. Eventually, I came to realize the duality of those perspectives would be exactly what made me who I was. I won’t shy away from that.”

“To me, ethnicity is more medically concrete than race. Ethnicity is your personal history, race is a construct used to discriminate by distributing power unequally.”

### Algorithmic Reform

The excerpts below are drawn from participant responses to “Do you have any additional thoughts or concerns to share?”. This question was posed after participants responded to questions about algorithmic reform based on three revised and one unrevised clinical calculators.

“The biggest concern I have is the exclusion of Africans. To simply group all Africans in ‘African-American’ will cause a lot of misdiagnosis as these people are very diverse and medical diagnosis cannot be grouped. I think being asked for your country and race and ethnicity is the only way to ensure fairness and more accurate diagnosis.”

“I understand why medical experts want to reduce harm and improve accuracy, but I still feel uneasy about using race or ethnicity in medical calculators because these categories can be imprecise and socially defined. I think it’s important to keep re-evaluating these tools and, where possible, move toward using direct biological or social factors rather than race itself.”

“I am aware that some racial groups are really more likely to have certain diseases like Diabetes. Pacific people, Asians and Hispanics have higher diabetic risk. While Whites have higher risk for skin cancer compared to other races. I think race-specific calculations must be applied depending on the disease or medical condition and careful scientific investigation must be done before implementing them.”

“I’m not particularly concerned of the fact that race/ethnicity could be included in the calculator if this inclusion is statistically relevant and improves the care given to the patients, but I would want for them to be tested against possible harmful biases.”

“The calculators should be based on the country and place that you lived in you might be Asian but you never went there you have been always in New York. If by race it’ll be unfair it should be where the person lives and what are the habits of this person.”

“I’m not qualified in the medical field, so I’d prefer to leave it to my doctors. I don’t mind, however, of the race, gender or any other trait is used in the statistical analysis; if there are actually some patterns, it’s okay to use them for the benefit of the people in my opinion.”

### Experiencing Racism in the Healthcare System

Survey participants who answered “Yes” to the question “Have you ever felt that you were treated unfairly in the healthcare system because of your race or ethnicity?”, were asked the following: “If you feel comfortable sharing, please briefly describe your experience. You may skip this question if you prefer not to answer.” The excerpts below are drawn from participant responses to this question.

“I remember a hospitalization case of my father in well known government hospital. We remain in queue for 5 long hours for some clinical tests but patients outside of our country and doctors relatives from foreign country got all facilities at instance. It really felt helpless because condition of my father was serious and no doctors or nurses were ready to support.”

“I have experienced situations in healthcare where my symptoms were minimized or not fully investigated. In one instance, a doctor assumed my risk for certain conditions was low based on my race and did not initially recommend further testing, even though I continued to have symptoms. This made me feel that decisions were being influenced by stereotypes rather than my individual health concerns.”

“I felt like I was treated unfairly during labour, like I was expected to endure more pain, plus even during the aftercare my pain was ignored.”

“I was in a hospital after a major loss of blood because of a very heavy menstrual cycle. The were having a difficult time finding a vein and had to stick me multiple times. I was very tired and in a lot of pain to then hear a nurse comment about how the man trying to find my vein should not be so gentle because black peoples skin is thicker. The number of times I’ve heard this is ridiculous.”

“Because of my middle eastern race, they considered higher chance of possibility of some bad characteristics or having some diseases.”

“I went to a clinic for a tooth ache once and it was really clear that they just wanted to pull it. They said would could save it, but it would be really expensive. as if I couldn’t afford it.”

“I feel like I have been to clinics where everyone shared the same fair complexion, and blonde hair & they were very rude to me. Made comments about they see weight struggles in Native Americans (I am not overweight, but they mentioned in passing).

“I consider myself Native American, but there is a lot of mixed blood going back a few generations. In my case, how I am perceived is dependent on the individual person doing the perceiving. Sometimes I am seen as White, sometimes Native, and sometimes they just look me up and down and say, in a very queer tone, ‘What are you?’

## Footnotes

1 GLI: Global Lung Function Initiative.

2 eGFR: estimated glomerular filtration rate; CKD-EPI: Chronic Kidney Disease Epidemiology Collaboration.

3 PREVENT: Predicting Risk of cardiovascular disease EVENTs.

4 FRAX: Fracture Risk Assessment Tool, maintained by this foundation.

