## Supplementary material for "Views on Race, Ethnicity, and Algorithmic Reform in Medicine: A Multinational Opt-In Survey": Questionnaire

### Informed Consent

THE UNIVERSITY OF BRITISH COLUMBIA
Faculty of Pharmaceutical Sciences

**Consent Form**

***Perspectives About Using Race or Ethnicity in Clinical Algorithms and Calculators***

**Who is conducting the study?**


Principal Investigator: Dr. Mohsen Sadatsafavi, MD PhD
Faculty of Pharmaceutical Sciences
University of British Columbia

Co-investigator: Amin Adibi, MSc
Faculty of Pharmaceutical Sciences
University of British Columbia

Additional Study Team Member: Xingyu Le
Faculty of Pharmaceutical Sciences
University of British Columbia


#### Who is funding this study?

The study is funded by UBC Public Scholars Initiative.

#### Purpose of the Study

Clinical algorithms and calculators are tools used to support clinical decision-making. The purpose of this study is to understand public opinion on using race or ethnicity in these calculators. You are invited because you are a registered Prolific survey taker and at least 18 years old.

#### What will happen if you participate?

If you consent, you will complete an online survey where you will review clinical calculators and answer questions about race or ethnicity in these tools. The survey should take about 15 minutes. You may skip any demographic or sensitive questions without explanation.

#### Risks of Participation

We do not think there is anything in the study that could harm you. Some questions may feel sensitive or personal. You may skip any of these questions you do not wish to answer.

#### Benefits of Participation

You may learn more about clinical calculators. Your input will help improve fairness in clinical calculators, which may benefit patients in the future.

**Privacy and Confidentiality**
Recruitment is through Prolific. Researchers only see your Prolific ID, not personal identifiers. Prolific does not store your survey responses. Data will be stored on secure, encrypted servers at UBC for at least five years. You may exit the survey at any time before completion. At the end of the survey, you will be asked to reconfirm your consent. If you choose not to submit at the end, your responses will not be recorded. Once the survey is submitted, withdrawal is not possible because responses are anonymous. This online survey is hosted by UBC Survey Tool (provided by Qualtrics). The survey data is kept secure and is stored and backed up in Canada and is subject to Canadian Law. If required by granting agencies or journals, de-identified data may be shared in open access repositories to help reviewers and other researchers verify findings and improve transparency and reproducibility.

**Compensation**
You will receive payment directly from Prolific as outlined in the survey invitation email, providing your response passes attention, authenticity, and quality checks.

**Use of Results**
Results will inform the design of fairer clinical calculators and may appear in a graduate thesis or academic publications. De-identified data may also be shared in open access repositories if required by journals or granting agencies.

**Voluntary Participation and Withdrawal**
Participation is voluntary. You may withdraw at any time before completing the survey or skip sensitive questions without explanation. Your decision will not affect your relationship with UBC or Prolific.

**Contact for Questions**

**Complaints or Concerns**
If you have concerns about your rights as a research participant and/or your experiences, contact the Research Participant Complaint Line in the UBC Office of Research Ethics at 604-822-8598, toll free 1-877-822-8598, or or call. Please reference the study number H25-02789 when contacting the Complaint Line so the staff can better assist you.

**Consent Statement**

By selecting 'I consent', you confirm that you have read and understood this form and agree to participate in this study.

Do you consent to participate in this survey?

- ○ I consent, begin the study
- ○ I do not consent, I do not wish to participate

**Prolific ID**

What is your Prolific ID?
*Please note that this response should auto-fill with the correct ID.*

${e://Field/PROLIFIC_PID}

**Past Experience**

Has a doctor or nurse ever told you that your race or ethnicity played a role in a medical decision or recommendation?

- ○ Yes
- ○ No
- ○ I am not sure

**Explainer / Transition**

Doctors sometimes use tools called **clinical algorithms or calculators**, to help make decisions about patient care. These tools take pieces of information—like someone's age, weight, blood pressure, or lab test results—and combine them to give an estimate. Some are simple formulas; others use more advanced methods like artificial intelligence (AI). For example, a calculator might predict how likely someone is to develop heart disease in the next ten years.

These predictions don't replace a doctor's judgment, but they can give useful information. Some calculators have also included race or ethnicity as one of the factors that go into the prediction. That means the results could come out differently depending on which racial or ethnic category a person chooses. This survey is about understanding how people feel about that practice.

In the following sections, you will see **four examples** of such calculators.

We ask you to **not** use AI chatbots (tools like ChatGPT or Claude) to answer this survey. We are interested in your personal opinion.

**Spirometry-Example**

#### Lung Function (Spirometry)

Spirometry is a breathing test that measures how well lungs work. It helps diagnose and monitor lung diseases. Doctors compare your results to reference values to decide if a patient's lungs are normal. This is similar to how doctors check a baby's growth against chart to make sure they baby is growing normally.

These normal values are predicted using factors like sex, height, age, and sometimes race or ethnicity. For example, some reference equations (such as GLI-2012) ask patients to identify as White, African American, Northeast Asian, Southeast Asian, or Other/Mixed.

If you were asked to identify with one of these categories for a lung function test, which one would you choose?

- ○ White
- ○ African American
- ○ Northeast Asian
- ○ Southeast Asian
- ○ Other/Mixed
- ○ Prefer not to answer

To what extent do you agree or disagree with the following statements?

| | Strongly disagree | Somewhat disagree | Neither agree nor disagree | Somewhat agree | Strongly agree |
|---|---|---|---|---|---|
| I feel represented by the race or ethnicity categories (as listed in the previous question) used to interpret lung function test results. | ○ | ○ | ○ | ○ | ○ |
| I believe that using race or ethnicity in spirometry could help ensure better care for me. | ○ | ○ | ○ | ○ | ○ |

| | Strongly disagree | Somewhat disagree | Neither agree nor disagree | Somewhat agree | Strongly agree |
|---|---|---|---|---|---|
| I worry that using my race or ethnicity in spirometry to support clinical decisions (for example, to assess the severity of lung diseases) might harm me. | ○ | ○ | ○ | ○ | ○ |

An alternative reference equation, called GLI-Global, does not ask for patient's race or ethnicity, and instead uses a single reference equation averaged across the four racial and ethnic groups (White, African American, Northeast Asian and Southeast Asian). Which of the following statements best describes your initial reaction to GLI-Global?

○ I feel more comfortable with the reference normal value does not depend on race and ethnicity.
○ I feel more comfortable when the reference normal value takes my race or ethnicity into account.
○ I feel neutral / unsure.
○ I prefer to self-describe (please specify): ______

**eGFR Example**

**Kidney Function**

Kidney function estimation (eGFR) is a calculation based on a blood test that checks how well your kidneys filter waste from your blood. It measures creatinine (a waste product from muscles) and considers factors like age and sex—and sometimes race or ethnicity. For example, the widely used 2009 CKD-EPI equation estimated higher kidney function for Black people.

The 2009 CKD-EPI kidney function equation required patients to be classified as either “Black” or “non-Black.” If you were asked to identify with one of these categories, which would you select?

○ Black
○ Non-Black
○ Prefer not to answer

To what extent do you agree or disagree with the following

statements?

| | Strongly disagree | Somewhat disagree | Neither agree nor disagree | Somewhat agree | Strongly agree |
|---|---|---|---|---|---|
| I feel represented by the race or ethnicity categories (as listed in the previous question) used to estimate kidney function. | ○ | ○ | ○ | ○ | ○ |
| I worry that using race or ethnicity in kidney function tests to support clinical decisions (for example, to evaluate the need for a kidney transplant) might harm me. | ○ | ○ | ○ | ○ | ○ |
| I believe that using race or ethnicity in kidney function estimation could help ensure better care for me. | ○ | ○ | ○ | ○ | ○ |

An alternative race-free method for estimating kidney function does not include an adjustment for Black patients. Which of the following statements best describes your initial reaction to the race-free equation?

- ○ I feel more comfortable when the equation does not consider my race or ethnicity.
- ○ I feel more comfortable when my race or ethnicity is considered.
- ○ I feel neutral / unsure.
- ○ I prefer to self-describe (please specify): ________

**CVD Example**

**Heart Disease Risk**

Heart disease risk calculators help doctors estimate a person's chance of developing heart disease, often over the next 10 years.

Some heart disease risk calculators ask for race or ethnicity. For example, ASCVD 2013 Risk Calculator from the American College of Cardiology (ACC) includes race categories: White, African American, or Other.

If you were asked to select a race or ethnicity category for heart disease risk calculator, which option would you choose?

- ○ White
- ○ African American
- ○ Other

○ Prefer not to answer

To what extent do you agree or disagree with the following statements?

| | Strongly disagree | Somewhat disagree | Neither agree nor disagree | Somewhat agree | Strongly agree |
|---|---|---|---|---|---|
| I feel represented by the race or ethnicity categories (as listed in the previous question) used to estimate heart disease risk. | ○ | ○ | ○ | ○ | ○ |
| I worry that using race or ethnicity in heart disease risk calculators to support clinical decisions (for example, to evaluate the need to go on blood pressure medication) might harm me. | ○ | ○ | ○ | ○ | ○ |
| I believe that using race or ethnicity in heart disease risk calculator could help ensure better care for me. | ○ | ○ | ○ | ○ | ○ |

An alternate heart risk calculator, the PREVENT score, does not ask for patient's race or ethnicity and instead optionally asks for the patient's postal or zip code. Which of the following statements best describes your initial reaction to PREVENT heart disease risk calculator?

○ I feel more comfortable when the equation uses postal or zip code instead of my race or ethnicity.
○ I feel more comfortable when my race or ethnicity is considered.
○ I feel neutral / unsure.
○ I prefer to self-describe (please specify): ________

**FRAX-example**

**Fracture Risk Assessment**

FRAX is a calculator that helps doctors estimate the patient's risk of breaking a bone in the next 10 years. It is mainly used to assess osteoporosis risk in older adults. FRAX asks for the patient's country. For some countries including US, Singapore, South Africa, and Malaysia, FRAX also asks for race or ethnicity.

If you were asked to select a country for FRAX fracture risk calculator, which option would you choose?

To what extent do you agree or disagree with the following statements?

| | Strongly disagree | Somewhat disagree | Neither agree nor disagree | Somewhat agree | Strongly agree |
|---|---|---|---|---|---|
| I feel represented by the country/race/ethnicity categories (as listed in the previous question) used to estimate bone fracture risk. | ○ | ○ | ○ | ○ | ○ |
| I worry that using country/race/ethnicity in FRAX calculator to support clinical decisions (for example, to evaluate the need to start osteoporosis treatment) might harm me. | ○ | ○ | ○ | ○ | ○ |
| I believe that using country/race/ethnicity in FRAX calculator could help ensure better care for me. | ○ | ○ | ○ | ○ | ○ |

**Reveal**

You reviewed four use cases for clinical calculators: Two of these help doctors decide if someone's lungs and kidneys perform normally, and two help predict the risk of heart disease and bone fracture. In recent years, some medical societies have re-evaluated how race or ethnicity is used in clinical calculators to ensure fairness to all.

In 2023, the top American and European medical societies for lung doctors recommended the lung function labs to **stop using race-specific references and switch to the race-averaged equations**. The experts were concerned that the race-specific references might make it more likely for doctors to miss lung disease in Black and Asian individuals. The race-averaged reference uses average values from White, African American, Northeast Asian, and Southeast Asian groups.

Does this official recommendation affect how you feel about using race or ethnicity to decide whether someone's lung function is normal?

- ○ Yes, knowing the official recommendation I now prefer the race-averaged approach
- ○ No, I prefer to form my own opinion regardless of official recommendations
- ○ No, I already preferred the race-averaged approach before learning this

○ No, I still have concerns about the race-averaged approach despite the recommendation

○ I'm not sure

In 2024, the International Osteoporosis Foundation recommended that FRAX bone fracture risk calculator **remain race-specific in the US.** The experts were concerned that a race-free calculator for bone fracture would unfairly discriminate against the Black, Asian and Hispanic communities in the US. FRAX asks for patients country, and in some countries asks for patients race or ethnicity.

Does this official recommendation affect how you feel about using race or ethnicity for predicting the risk of bone fracture?

○ Yes, knowing the official recommendation I now prefer race-specific risk prediction for bone fractures

○ No, I prefer to form my own opinion regardless of official recommendations

○ No, I already preferred the race-specific approach before learning this

○ No, I still have concerns about the race-specific approach despite the recommendation

○ I'm not sure

Do you have any additional thoughts or concerns to share?

**Future Directions**

In your own words, what does "race or ethnicity" mean to you? *There are no right or wrong answers - we are interested in your personal perspective.*

Who do you think should have input into decisions about whether or how race or ethnicity is used in clinical calculators? **Select all that apply**.

☐ Government or public health agencies

☐ The general public

☐ Patient advocacy groups or organizations

☐ Individual patients (for decisions about their own care)

☐ Medical researchers and scientists

☐ Doctors and healthcare providers

☐ Other (please specify) ____________

Imagine a doctor is using a clinical calculator to determine your risk for a type of cancer. How comfortable are you with the calculator using each of the following pieces of information?

| | Extremely uncomfortable | Somewhat uncomfortable | Neither comfortable nor uncomfortable | Somewhat comfortable | Extremely comfortable |
|---|---|---|---|---|---|
| Your home postal (or zip) code | ○ | ○ | ○ | ○ | ○ |
| Your race or ethnicity | ○ | ○ | ○ | ○ | ○ |
| Your genetic ancestry | ○ | ○ | ○ | ○ | ○ |
| Your environmental exposures (e.g. air pollution) | ○ | ○ | ○ | ○ | ○ |
| Your biological sex | ○ | ○ | ○ | ○ | ○ |
| Your country of origin | ○ | ○ | ○ | ○ | ○ |
| Your age | ○ | ○ | ○ | ○ | ○ |

Clinical algorithms can range from simple formulas to complex AI systems. Would your concerns about using race or ethnicity differ depending on the type of tool?

○ I would be more concerned about race or ethnicity in AI systems than in simple formulas.
○ I would be equally concerned regardless the type of tool.
○ I would be less concerned about race or ethnicity in AI systems than in simple formulas.
○ I'm not concerned about race or ethnicity being used in any tools.
○ I'm not sure.

**Replication**

To what extent do you agree or disagree with the following statements?

| | Strongly disagree | Somewhat disagree | Neither agree nor disagree | Somewhat agree | Strongly agree |
|---|---|---|---|---|---|
| My doctor should use my race or ethnicity in clinical care as long as my doctor believes it is best for my care. | ○ | ○ | ○ | ○ | ○ |
| There are no circumstances under which a doctor should use my race or ethnicity in clinical care. | ○ | ○ | ○ | ○ | ○ |

| | Strongly disagree | Somewhat disagree | Neither agree nor disagree | Somewhat agree | Strongly agree |
|---|---|---|---|---|---|
| I am comfortable with my doctor using my race or ethnicity in clinical care as long as it is recommended by practice guidelines. | ○ | ○ | ○ | ○ | ○ |
| I am comfortable with my doctor using my race or ethnicity in clinical care if they talk to me about how and why they are using it. | ○ | ○ | ○ | ○ | ○ |

When you are seeking healthcare, how often are you asked what race or ethnicity group you belong to (either by the doctor or on a form)?

○ Never
○ Sometimes
○ About half the time
○ Most of the time
○ Always

Have you ever had the experience of noticing that a doctor, nurse, or clinic staff have filled out your race or ethnicity on a form or computer screen without asking you?

○ Yes, and it matched my own choice
○ Yes, and it did not match my own choice
○ No
○ Not sure

In general, do you think you would receive better or worse care from doctors or health care providers who share your racial and ethnic background, or would it not make much difference?

○ Would receive better care
○ Would receive worse care
○ Wouldn't make much difference

In health and medicine, how much of a problem is bias and unfair treatment based on patients' race or ethnicity?

○ A major problem
○ A minor problem
○ Not a problem
○ Unsure

**Racism**

Have you ever felt that you were treated unfairly in the healthcare system because of your race or ethnicity?

- ○ Yes
- ○ No
- ○ Unsure
- ○ Prefer not to say

If you feel comfortable sharing, please briefly describe your experience.
*You may skip this question if you prefer not to answer.*

**Demographic**

The following section asks some demographic questions to help us better understand the diversity of our participants and interpret the survey results.

All demographic questions are optional. Please select "Prefer not to say" for any questions you are not comfortable answering.

Do you have a background in healthcare or health sciences (through work, study, or training)?

- ○ Yes
- ○ No
- ○ Prefer not to say

Which population group or groups best describe you? Select all groups that apply, or you may self-describe if preferred.

- ☐ African (Central, East, West, or Southern Africa)
- ☐ African American
- ☐ Caribbean
- ☐ East Asian
- ☐ Hispanic or Latina/Latino
- ☐ Indigenous

☐ Middle Eastern and North African
☐ South Asian
☐ Southeast Asian
☐ White
☐ [ ] I prefer to self-describe (please specify)
☐ Prefer not to say

**Final Step: Please review and confirm your participation**

Your responses will help researchers understand public opinion about race or ethnicity in clinical calculators. Findings will inform design of clinical calculators to promote fairness and equity in healthcare. All responses are anonymous. If you have questions, contact. For concerns about your rights as a participant, contact the UBC Office of Research Ethics at or call +1-877-822-8598 (toll-free). Please reference REB# H25-02789.

Would you like to submit your survey results?

○ Yes, submit my responses
○ No, I want to revoke my consent

Powered by Qualtrics
